# Improving Tuberculosis Control: Assessing the Value of Medical Masks and Case Detection – A Multi-Country Study with Cost-Effectiveness Analysis

**DOI:** 10.1101/2024.06.18.24309075

**Authors:** Dipo Aldila, Basyar Lauzha Fardian, Chidozie Williams Chukwu, Muhamad Hifzhudin Noor Aziz, Putri Zahra Kamalia

## Abstract

Tuberculosis (TB) remains a significant global health concern, necessitating effective control strategies. This paper presents a mathematical model to evaluate the comparative effectiveness of medical mask usage and case detection in TB control. The model is constructed as a system of ordinary differential equations and incorporates crucial aspects of TB dynamics, including slow-fast progression, medical mask utilization, case detection, treatment interventions, and differentiation between symptomatic and asymptomatic cases. A key objective of TB control is to ensure that the reproduction number, ℛ_*c*_, remains below unity to achieve TB elimination or persistence if ℛ_*c*_ exceeds one. Our mathematical analysis reveals the presence of a transcritical bifurcation when the ℛ_*c*_ = 1 signifies a critical juncture in TB control strategies. These results confirm that the effectiveness of case detection in diminishing the endemic population of symptomatic individuals within a TB-endemic equilibrium depends on exceeding a critical threshold value. Furthermore, our model is calibrated using TB yearly case incidence data per 100,000 population from Indonesia, India, Lesotho, and Angola, and we employ the Bootstrap Resampling Residual Approach to assess the uncertainty inherent in our parameter estimates and provide a comprehensive distribution of parameter values. Despite a declining trend in new incidence, these four countries exhibit a reproduction number greater than one, indicating persistent TB cases in the presence of ongoing TB control programs. We employ the Partial Rank Correlation Coefficient (PRCC) in conjunction with the Latin Hypercube Sampling (LHS) method to conduct global sensitivity analysis of the *ℛ*_*c*_ parameter for each fitted parameter in every country. We find that the medical mask use is more sensitive to reduce ℛ_*c*_ compared to the case detection implementation. To further gain insight into the necessary control strategy, we formulated an optimal control and studied the cost-effectiveness analysis of our model to investigate the impact of case detection and medical mask use as control measures in TB spread. Cost-effectiveness analysis demonstrates that combining these interventions emerges as the most cost-effective strategy for TB control. Our findings highlight the critical importance of medical masks and their efficacy coupled with case detection in shaping TB control dynamics, elucidating the primary parameter of concern for managing the control reproduction number. We envisage our findings to have implications and be vital for TB control if implemented by policymakers and healthcare practitioners involved in TB control efforts.

## 1. Tuberculosis Recent Facts

Tuberculosis (TB) is an infectious disease caused by the bacterium *Mycobacterium tuberculosis* [1]. It primarily affects the lungs but can also target other parts of the body, such as the kidneys, spine, and brain. TB is spread through the air when an infected person coughs, sneezes, or talks, releasing tiny droplets containing the bacteria [2]. The infection usually presents in two forms: latent TB infection (LTBI) and active TB disease [3]. In LTBI, the bacteria remain dormant within the body, and the infected individual does not experience symptoms or feel sick. However, they are at risk of developing active TB if their immune system weakens. Active TB, on the other hand, leads to noticeable symptoms like persistent cough, chest pain, fatigue, fever, night sweats, and weight loss. It is essential to diagnose and treat active TB promptly, as it can be life-threatening if left untreated.

Preventive measures, such as vaccination (with the Bacillus Calmette-Guérin vaccine) for children and early identification and treatment of infected individuals, are crucial in controlling the spread of TB and reducing its impact on public health [4]. Effective treatment of TB involves a combination of antibiotics taken over a specific period, usually six to nine months, to ensure complete eradication of the bacteria and reduce the risk of drug resistance [5]. In some cases of drug-resistant TB, treatment may require a more extended and challenging regimen. Medical masks, such as surgical masks or N95 respirators, can provide some level of protection against the transmission of TB, but they are not specifically designed as a primary preventive measure for TB [6]. TB is primarily spread through the air when an infected person coughs, sneezes, or talks, releasing infectious droplets that can be inhaled by others nearby. While medical masks can help reduce the risk of inhaling large respiratory droplets that contain the TB bacteria, they are not entirely effective in preventing transmission.

The use of mathematical models to aid scientists in understanding the mechanisms of disease spread on a population scale has a long history. Many of these models have been inspired by the famous epidemic model developed by Kermack and McKendrick [7]. Subsequently, numerous mathematical models have been introduced to enhance our understanding of the spread of various well-known diseases, such as dengue [8, 9], malaria [10, 11], HIV/AIDS [12], pneumonia [13], COVID-19 [14, 15], tuberculosis [16, 17], and many more. Recently, several mathematical models have been developed to gain a more specific understanding of the mechanisms underlying tuberculosis transmission on a population scale. Authors in [18] studied the impact of isolation for TB cases in India using their SIQR model, in which they predicted that isolating half of the multi-drug-resistant TB (MDR-TB) cases could lead to a substantial reduction in TB incidence by 2025, with a concurrent decline in estimated MDR-TB incidence. A mathematical model considering treatment introduced by authors in [19]. Their parameter values were estimated using incidence data from Pakistan. They emphasized the significance of reducing the basic reproduction number to less than one as a pivotal strategy for eradicating TB epidemics and underscored the need to decrease treatment failure cases to reduce TB infectivity. The study also highlighted the effectiveness of isolating infective individuals through the reduction of the transmission coefficient. Okuonghae 2022 [20] performed a thorough mathematical analysis on a simplified stochastic TB disease model with case detection. The author found that the only disease persistence depends on the case detection parameter. Disease eradication showed an independent relation with the case detection parameter. With the existence of pharmaceutical intervention in controlling the spread of TB, [21] proposed a stochastic TB model by incorporating the effect of antibiotic resistance. They found sufficient conditions (dependency between parameters on the reproduction number) for the extinction of TB from the population. An age-structured model for TB dynamics was constructed by authors in [22]. It was found that the detection of LTBI could increase or decrease the reproduction number depending on the model parameter condition.

There are many options to prevent tuberculosis infection, with the most popular method being the use of vaccines. In a study by the authors in [23], vaccination aspects were incorporated into their delay-differential equation model. The authors discovered that there is a minimum number of vaccinations such that vaccine intervention could effectively suppress the spread of TB. A different approach, as demonstrated by the authors in [24], involves the use of an optimal control approach to model the impact of vaccines and treatments on TB dynamics. Besides vaccines, the intervention of treatment is also important for TB control programs. The authors in [25] considered the impact of treatment with three different latently infected classes in their model. Furthermore, an innovative fractional-order stochastic differential equation model was introduced by the author in [ 26] to analyze the impact of treatment on TB.

Despite these efforts, it is worth noting that not many mathematical models have considered the use of medical masks and case detection, similar to the strategies used during the COVID-19 pandemic, as simple and easy-to-implement prevention strategies for TB. Therefore, our proposed model in this article will incorporate two different interventions for preventing and controlling the spread of TB, namely the use of medical masks and case detection. The model developed using a five-dimensional system of ordinary differential equations. Mathematical analysis and numerical experiments conducted to demonstrate the long-term behaviour of the proposed model.

The layout of our article is organized into several key sections, each addressing distinct aspects of our research. In Section 1, we provide an overview of recent facts about TB and review previous mathematical models relevant to our study. Section 2 outlines our model assumptions, construction methodology, and details the parameter estimation process. In Section 3, we delve into the dynamical analysis of our model, focusing on equilibrium points, the reproduction number, and bifurcation analysis to understand the underlying TB dynamics of our model. Moving forward, Section 4 is dedicated to global sensitivity analysis, where we explore the impact of medical masks and case detection on controlling the reproduction number of TB. In Section 5, we present the results of our optimal control simulations and conduct a cost-effectiveness analysis to evaluate the efficacy and efficiency of various TB control strategies. Section 6 encapsulates our conclusions, summarizing key findings, implications, and a venues f or future research. Additionally, we provide appendices containing proofs and visualizations of theorems presented throughout the manuscript, offering supplementary information to enhance the understanding and rigor of our study.

## 2. Mathematical model construction and parameter estimation

### 2.1. Model construction

Let the human population be divided into five compartments, namely susceptible (*S*), exposed/latent (*E*), infected asymptomatic, undetected and untreated (*I*_1_), infected symptomatic, detected and treated (*I*_2_), and recovered (*R*). Therefore, the total human population is given by:

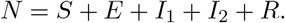

To construct the mathematical model for tuberculosis transmission in this research, several important assumptions need to be declared first.

1. **Case detection**. Case detection is a crucial component of tuberculosis (TB) control efforts aimed at identifying and diagnosing individuals with active TB disease. Effective case detection is essential for initiating prompt treatment, preventing further spread of the disease, and ultimately reducing the burden of TB [27]. Based on this importance, we include the case detection effort (*u*_1_) in our model to find symptomatic undetected individuals in the field. Hence, *u*_1_*I*_1_ represents the newly detected symptomatic TB-infected individuals.
2. **Effect of medical mask use**. With the escalation of respiratory diseases, including tuberculosis (TB), the utilization of medical masks emerges as one of the most prevalent and pragmatic non-pharmaceutical interventions aimed at diminishing the risk of infection and disease transmission. While the effectiveness of wearing medical masks in combating diseases remains a topic of debate [28, 29, 30], numerous pre-COVID-19 pandemic studies highlight the potential efficacy of medical mask usage among TB-active patients in curtailing the spread of tuberculosis. In three human studies conducted in healthcare settings, a reduction in TB cases was observed among the participants who used the medical masks [31, 32, 33]. The findings were also consistent with an animal study by Dharmadhikari et al. [34]. The study reported that 56% decreased risk of TB transmission in a group of guinea pigs when exposed to air from active TB patients who wore masks. To model the impact of medical mask use, let us denote *β* and *u*_2_ as the TB successful infection rate and rate of medical mask use, respectively. Furthermore, it is assumed that individuals in compartment *I*_2_ cannot transmit TB to others because they are presumed to adhere to recommendations to reduce close contact with others, either by maintaining distance, isolation, or quarantine. Hence, we have *u*_2_*I*_1_ represent the proportion of infected individuals who use medical masks, while (1 − *u*_2_)*I*_1_ are those who do not. Hence, the total new infection caused by infected individuals who do not use medical masks is given by *β*(1 − *u*_2_)*I*_1_*S*. Further, we assume that the use of a medical mask may reduce the successful infection rate with an efficacy of *ξ*. The more effective the medical mask, the larger the value of *ξ*. Hence, the total number of new infections caused by infected individuals who use medical masks is given by (1 − *ξ*)*βu*_2_*I*_1_*S*. Therefore, the total number of new infections, Λ(*S, I*_1_) caused by *I*_1_ is given by:

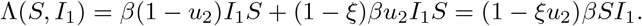
3. **Slow-fast progression**. TB infection can exhibit different progression patterns, including slow and fast progression [35]. In slow progression, the infection advances gradually over an extended period. Individuals with slow-progressing TB may not show noticeable symptoms for a significant period after being exposed to the bacteria. Latent TB may later progress to active TB disease under certain conditions, such as a weakened immune system. On the other hand, fast progression refers to a more rapid development of active TB disease after exposure to the bacteria. Individuals with fast-progressing TB may experience symptoms relatively soon after being infected. Based on this, we assume that the total number of new infections given by Λ(*S, I*_1_) may experience slow progression with a probability of *p* or fast progression with a probability of *q*. Note that *p* + *q* = 1. Hence, the proportion of newly infected individuals who experience slow progression is given by:

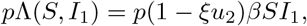

while for fast progression is given by

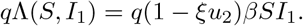

Using the above assumptions, the model construction is given as follows: We assume that the recruitment rate is always constant with a value of *δ*. Under the impact of medical mask use, the number of susceptible individuals may decrease due to a new infection by *I*_1_ individual, given by (1−*ξu*_2_)*βSI*_1_. Due to slow-fast progression, a proportion of newly infected individuals will experience slow-progression (*p*(1 − *ξu*_2_)*βSI*_1_) and enter the *E* compartment, while the rest will experience fast-progression (*q*(1 − *ξu*_2_)*βSI*_1_) and move to the *I*_1_ compartment. Furthermore, it is assumed that latent individuals can undergo an increase in infection status, becoming infectious and exhibiting symptoms, thus requiring treatment. We use *ϵ* to represent this phenomenon, which allows the transition from *E* to *I*_2_. Without any early case detection, latent individuals will experience TB progression and become infected. Hence, there is a transition rate from *E* to *I*_1_ due to infection progression, denoted by *θ*.

Due to case detection for the symptomatic individual *I*_1_, we have a transition from *I*_1_ to *I*_2_ with a rate of *u*_1_. Both *I*_1_ and *I*_2_ may recover from TB with a rate of recovery given by *k*_1_ and *k*_2_, respectively. Since *I*_2_ gets an intensive treatment, we have *k*_2_ *> k*_1_. In addition to natural death occurring at a rate *µ* in each compartment, there is a death rate specifically due to TB for compartments *I*_1_ and *I*_2_ with rates *d*_1_ and *d*_2_, respectively.

Hence, the TB model with interventions such as case detection and medical masks is given by:

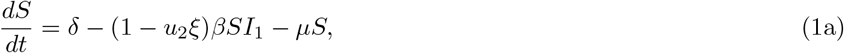

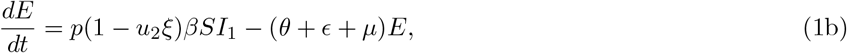

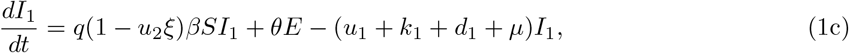

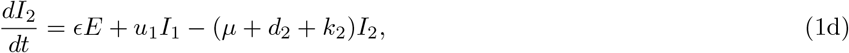

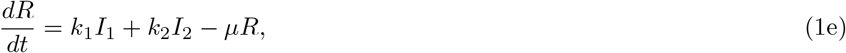

with non-negative initial conditions. Let

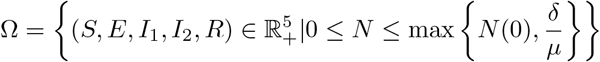

defined as the invariant region for the model (1). As long as the initial conditions are in Ω, the solution of the above system will always remain in Ω. Interested readers can see Appendix A for the proof.

### 2.2. Estimating Model Parameters Using Data Fitting

Before conducting simulations for the optimal control problem in Section 6, we performed parameter estimation for our model using yearly new incidence data per 100,000 individuals from four different countries: Indonesia, India, Angola, and Lesotho. The data, starting from 2000 to 2020, were obtained from [36].

We aimed to find the best-fit parameter and best-fit initial conditions of our model such that the Euclidean distance between the incidence data and model output simulation was minimized. Since the data is the yearly new incidence data, we fitted the data with the newly detected incidence of TB both from *E* and *I*_1_ compartments, i.e., *ϵE* and *u*_1_*I*_1_. Particularly, the following cost function was minimized

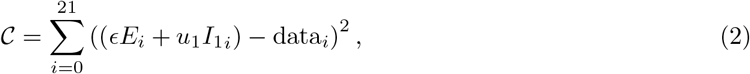

where 21 is the number of data points collected from the years 2000 to 2020 for each country. Other parameters were held constant as follows:

- The recruitment rate (*δ*) and the natural death rate (*µ*): From equation (1), the dynamic of the total human population is given by

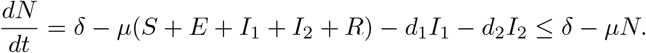 Hence, if we assume that *d*_1_ and *d*_2_ are relatively small, then the total population can be assumed to be constant. Hence, we have *δ* = *µN*. Given that the incidence data utilized for parameter estimation is presented as the incidence rate per 100,000 people [36], it follows that our population size, denoted as *N*, is equivalent to 100,000. Furthermore, since the average human life expectation is between 66.8 years in 2000 to 73.4 in 2019 [37], then we assume that 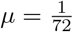.
- Medical mask efficacy (*ξ*): The use of surgical face masks on patients with MDR-TB has demonstrated a significant reduction in transmission, providing an additional measure to mitigate the spread of TB from infectious individuals [34]. Hence, from the same reference, we choose *ξ* = 56%.
- The TB latent progression to active TB (*θ*): Reactivation is the transition of a subclinical latent infection into active TB disease. Consequently, individuals with latent TB infection (LTBI) serve as a significant reservoir for the emergence of new active TB cases. According to [38], it takes approximately 2 years for an individual to progress from latent TB to active TB. Hence, we assume 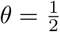.
- The recovery rate (*k*_1_ and *k*_2_): Most individuals with TB disease typically require a minimum of 6-12 months of TB treatment for a complete cure [39]. Hence, we assume *k*_2_ = 1. Since *k*_1_ *< k*_2_, then we assume *k*_1_ = 0.5.
- Death rate due to TB (*d*_1_ and *d*_2_): It is assumed that the death rate due to TB for *I*_1_ and *I*_2_ is 0.01.

The other parameters, namely the infection rate *β*, the proportion of slow-fast progression *p* and *q*, case detection rate *u*_1_, the proportion of medical mask use *u*_2_, and transition to symptomatic from latent individual *ϵ*, were estimated together with the initial conditions. Mathematically, it can be written as

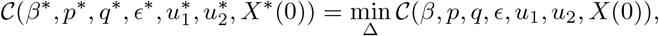

where *X*(0) is the set of initial condition of system (9), and Δ is the set of admissible range of parameter values. In this study, we employ the `fmincon` toolbox to estimate the parameters of our model, as described by equation (1). `fmincon` is a powerful optimization tool in MATLAB, typically utilized for solving constrained nonlinear optimization problems. In our adaptation of `fmincon` for ordinary differential equations (ODEs), we formulate the parameter estimation task as an optimization problem where the objective function represents the discrepancy between model predictions and observed data, subject to any pertinent constraints. We iteratively refine the model parameters by optimizing this objective function until a satisfactory fit to the data is achieved. Additionally, to assess the uncertainty inherent in our parameter estimates and provide a comprehensive distribution of parameter values, we employ the Bootstrap Resampling Residual Approach [40, 41] across all estimation results for four distinct countries. This approach allows us to generate multiple parameter sets by resampling residuals, providing insight into the variability and robustness of our model across different datasets and scenarios. The fitting results for the incidence data of four different countries are given in Figure 1, and the best-fit parameter values are listed in Table 1.

**Table 1:** The best-fit parameters and best-fit initial conditions for the fitted curves in Figure 1.

| Country | $\beta(10^{-5})$ | $p$ | $q$ | $\epsilon$ | $u_1$ | $u_2$ | $S(0)$ | $E(0)$ | $I_1(0)$ | $I_2(0)$ | $R(0)$ | $\mathcal{R}_0$ |
| --- | --- | --- | --- | --- | --- | --- | --- | --- | --- | --- | --- | --- |
| Indonesia | 3.356 | 0.885 | 0.115 | 0.121 | 0.151 | 0.5 | 34 152 | 1 546 | 1 208 | 2 179 | 57 | 2.907 |
| India | 1.752 | 0.899 | 0.101 | 0.087 | 0.102 | 0.5 | 58 975 | 2 075 | 1 549 | 5 543 | 523 | 1.711 |
| Lesotho | 2.061 | 0.883 | 0.117 | 0.155 | 0.198 | 0.5 | 80 871 | 1 586 | 2 921 | 3 012 | 475 | 1.967 |
| Angola | 1.251 | 0.898 | 0.102 | 0.045 | 0.13 | 0.5 | 93 995 | 1 821 | 1 649 | 3 027 | 621 | 1.247 |

**Figure 1:**
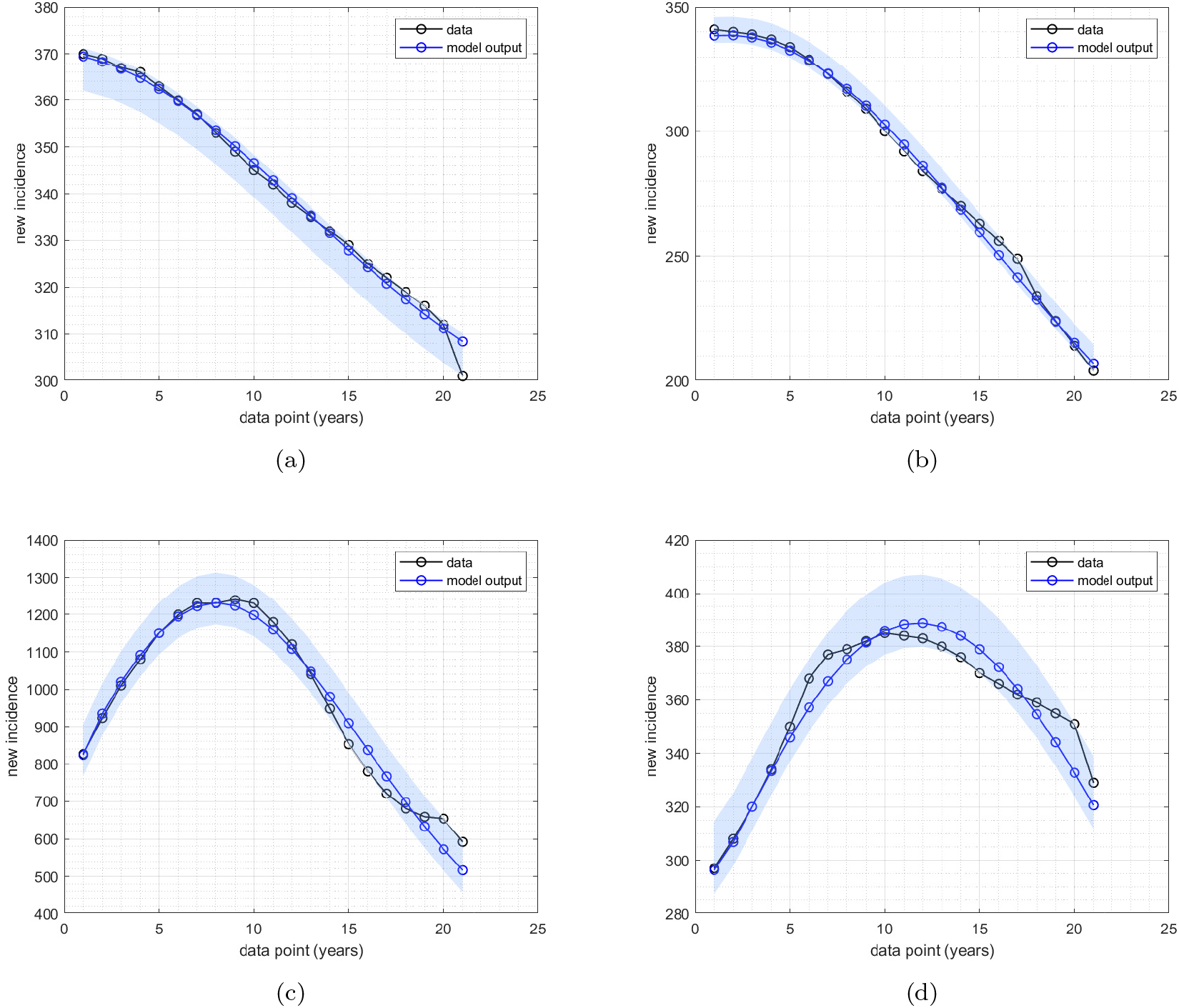
Fitted data of new incidence cases for (a) Indonesia, (b) India, (c) Lesotho, and (d) Angola. The solid black and blue lines represent the real data and simulated data, respectively.

In the next section, we provide a complete mathematical analysis of the model, including the existence and local stability of the equilibrium points of system (1)as well as the control reproduction number (ℛ_*c*_).

## 3. Dynamical analysis

### 3.1. *Disease-free equilibrium point* (ℰ_1_)

The disease-free equilibrium of system (1) was obtained by letting the right-hand side of system (1) to zero and setting *I*_1_ = *I*_2_ = 0. Hence, we have:

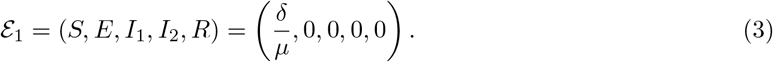

### 3.2. Control reproduction number (ℛ_c_)

#### Theorem 1.

*The control reproduction number of system* (1), *denoted by ℛ*_*c*_ *is given by:*

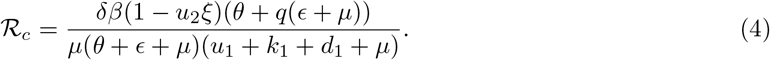

Please see (Appendix B) for the derivation of the control reproduction number.

It is common in many epidemiological models [42, 43, 44] for the reproduction number to determine whether the disease may die out or persist in the population. In many cases, authors have found that the disease can go extinct if the reproduction number is less than one, and it always has a chance to persist if the basic reproduction number is larger than one. In the next theorem, we use the results by authors in [45] to show the local stability criteria of the equilibrium ℰ_1_.

#### Theorem 2.

*The disease-free equilibrium* ℰ_1_ *of system* (1) *is locally asymptotically stable if ℛ*_*c*_ *<* 1, *and unstable if ℛ*_*c*_ *>* 1.

See (Appendix C) for the complete proof of Theorem 2

### 3.3. Global stability of the DFE

Using the approach in [46], we prove the existence of global asymptotic stability (GAS) for the disease-free equilibrium of our TB model. First, we rewrite equation (1) as follows:

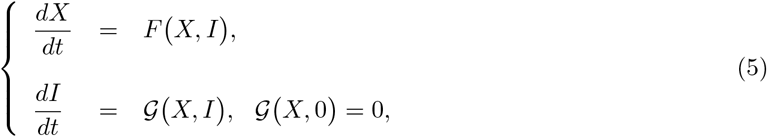

in which *X* = (*S, R)* ∈ ℝ^2^ and *I* = (*E, I*_1_, *I*_2_) ∈ ℝ^3^. Note that the variables *X* and *I* represents un-infectious and infectious TB-individuals respectively. For the model to be GAS at ℰ_1_, it needs to satisfy the following conditions as stated in [46], that is:

𝒞_1_) Local stability is guaranteed at *E* ^0^ whenever *R*_0_ *<* 1. *dX*
𝒞_2_) At 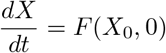 the DFE is globally asymptotically stable.
𝒞_3_) 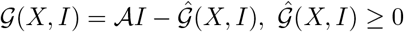 for (*X, I*) ∈ Ω, where *X*_0_ = ℰ_1_, *𝒜* = 𝒟_*I*_ *𝒢* (ℰ_0_) is a Metzler matrix, and Ω is our TB-model biologically feasible region.

#### Theorem 3.

*Let t >* 0, *then the disease-free equilibrium ℰ*_1_ *is GAS stable if ℛ*_0_.

See Appendix D for the complete proof of Theorem 3.

### 3.4. Endemic equilibrium point

Taking the right-hand side of system (1) equal to 0 and solving it with respect to each variable, then we have the endemic equilibrium point of system (1) given by:

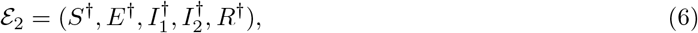

where

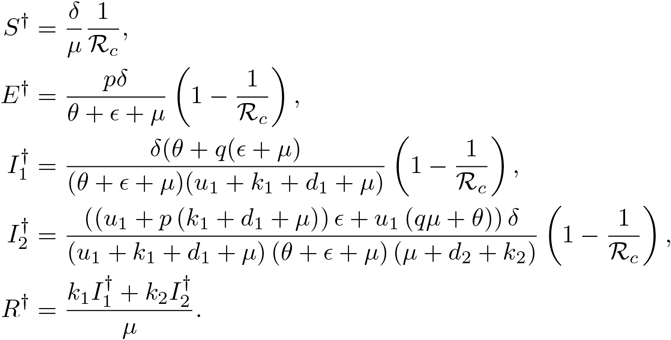

Based on the expression of ℰ_2_ above, we have the following theorem.

#### Theorem 4.

*There always exists a unique endemic-equilibrium point ℰ*_2_ *of system* (1) *if ℛ*_*c*_ *>* 1.

Proof. The proof of this theorem can be directly seen from the expressions of 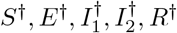. Each of these expressions should be positive. For any positive parameters, we will always have *S*^*†*^ *>* 0. On the other hand, 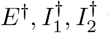 will be positive only if ℛ_*c*_ *>* 1. Lastly, *R*^*†*^ is always positive since the total population is less than 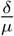. Hence, the proof is completed.

### 3.5. Non-existence of backward bifurcation

#### Theorem 5.

*System* (1) *always exhibits a transcritical bifurcation at ℛ*_*c*_ = 1.

We use Castillo-Song bifurcation theorem [47] to proof Theorem 5. See (Appendix E) for the complete proof of Theorem (5).

Based on Theorem 5, we can observe that the backward bifurcation phenomenon never occurs in our proposed TB model as described in (1). On the other hand, as per Theorem 3, we know that the disease-free equilibrium is globally asymptotically stable when ℛ_*c*_ *<* 1. Therefore, it is reasonable to hypothesize that the endemic equilibrium point is globally asymptotically stable when ℛ_*c*_ *>* 1. We leave the proof of this statement as an open problem for readers who may be interested.

### 3.6. Impact of medical mask and case detection on the endemic of TB

#### Comparison of the control and the basic reproduction number

In a simple case of no control intervention *u*_1_ = 0, *u*_2_ = 0, then we can reduce the control reproduction *ℛ*_*c*_ in the following basic reproduction number:

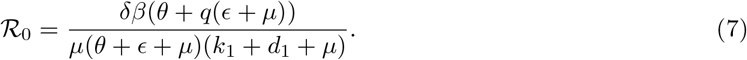

Since for all positive parameters, we have

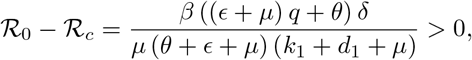

we have the following remarks.

##### Remark 1.

*Any positive intervention of medical mask use or case detection will always be successful in reducing the basic reproduction number ℛ*_0_.

#### Impact of medical mask and case detection to ℛ_c_

The expression of ℛ_*c*_ can be expressed as a function of ℛ_0_ as follows:

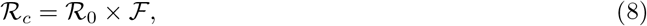

where 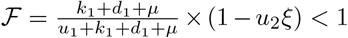 is the reduction factor of ℛ_0_ due to case detection *u*_1_ and medical mask use *u*_2_. Based on this, we have the following remark.

##### Remark 2.

*The following remark is the direct interpretation of expression in* (8)

1. *For a special case where the case detection rate tends to* ∞, *then we have:*

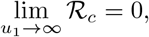

*which implies a massive intervention in case detection can reduce the reproduction number significantly*.
2. *For a special case when all individuals use medical mask* (*u*_2_ = 1), *then we have:*

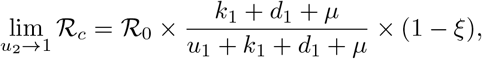

*which implies that using higher quality medical masks* (*ξ* → 1) *can lead to a more efficient reduction of the control reproduction number*.

#### Impact of case detection to 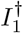 and 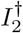

To analyze the impact of case detection and medical mask use on the size of *I*_1_ and *I*_2_ at the endemic equilibrium, we differentiate 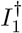 and 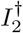 with respect to *u*_1_ and *u*_2_. Derivation of 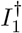 respect to *u*_1_ and *u*_2_ gives:

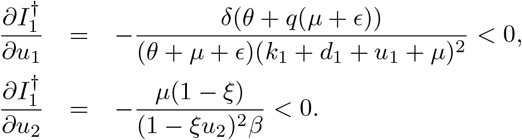

For any value of *u*_1_ *>* 0 and *u*_2_ ∈ [0, 1], the signs of 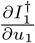 and 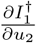 are always negative. This indicates that the size of 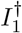 will always reduce whenever case detection or medical mask use is implemented for TB control. Furthermore, since *u*_1_ only appears in the denominator, both *u*_1_ and *u*_2_ have a significant impact on the change in the size of 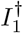 for an early implementation, as 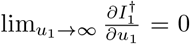 and 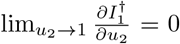. With these results, we have the following remark.

##### Remark 3.

*Implementation of case detection is always successful in reducing the number of asymptomatic infected individuals at the TB-endemic equilibrium*.

On the other hand, we have

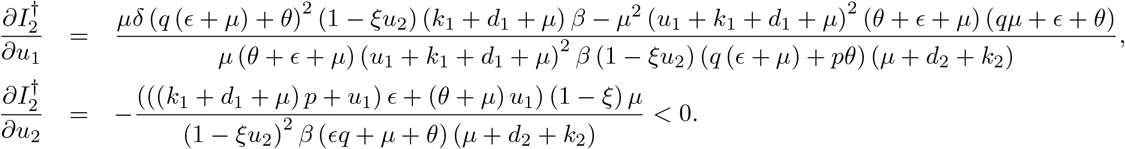

We can see that 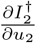 is always negative. Therefore, increasing medical mask use in the population will reduce the size of 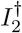. However, 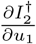 is not always negative. Solving 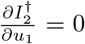 with respect to *u*_1_ gives us 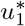 as a critical value at which the sign of 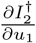 changes from positive to negative. With these results, we have the following remark.

##### Remark 4.

*The implementation of medical masks consistently reduces the number of symptomatic infected individuals. Additionally, there exists a critical value for case detection, beyond which the implementation of case detection successfully reduces the number of symptomatic infected individuals in the context of TB endemic equilibrium*.

The illustration of the aforementioned remarks can be seen in Figure 2 using estimation results for Lesotho (panel (a)) and for Indonesia’s data (panel (b)). In both panels, it is observed that there exists a minimum value 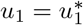 such that only when 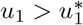 an increase in the case detection rate can reduce the number of individuals symptomatic with TB (*I*_2_) at the endemic equilibrium. Conversely, if 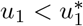, an increase in the case detection rate raises the value of *I*_2_ at the endemic equilibrium. Furthermore, *P*_2_ represents the value of *u*_1_ such that ℛ_0_ = 1. Therefore, for the case using Lesotho’s data, it is evident that the implementation of case detection can be relied upon to eliminate TB cases in Lesotho, specifically when *u*_1_ *>* 0.896. On the other hand, for the data from Indonesia, it is observed that there is no point *P*_2_ within the range of *u*_2_ ∈ [0, 1]. This implies that the case detection intervention cannot eliminate TB in Indonesia.

**Figure 2:**
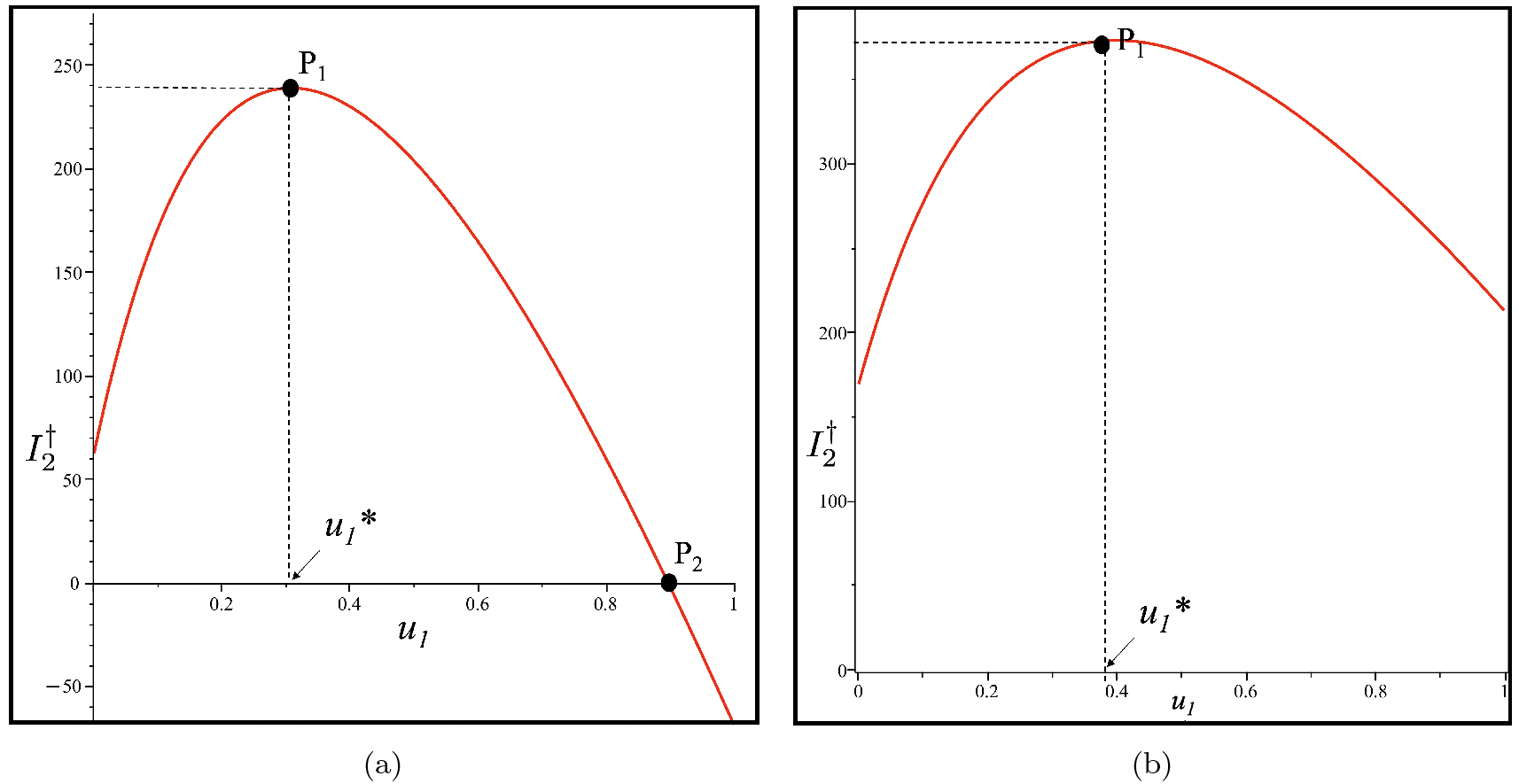
Curve of 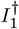 as a function of *u*_1_ using Lesotho and Indonesia data in panels (a) and (b), respectively. *P*_1_ is the turning point of 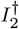, while *P*_2_ is *u*_2_ when *ℛ*_*c*_ = 1. When 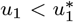, then 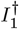 increases as *u*_1_ increases. The critical value 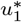 for Lesotho and Indonesia is 0.308 and 0.398, respectively. TB endemic equilibrium only exists when *u*_1_ *< P*_2_.

In this chapter, we have discussed the dynamic properties of the model presented in Equation (1). These dynamic properties include that the model (1) always has a stable TB-free equilibrium point for ℛ_*c*_ *<* 1, and it becomes unstable when ℛ_*c*_ *>* 1. The point ℛ_*c*_ = 1 serves as a bifurcation point where the stability of the TB-free equilibrium point changes, marking the emergence of the endemic equilibrium point. A new TB-endemic equilibrium point emerges and is always stable under conditions where ℛ_*c*_ *>* 1. For further visualization of these results, such as bifurcation diagrams and one-parameter sensitivity analysis, readers can refer to Section Appendix F.

In the following chapter, we will provide a study on parameter sensitivity, including global sensitivity analysis using the Partial Rank Correlation Coefficient (PRCC) and Latin Hypercube Sampling (LHS), as well as two-parameter sensitivity analysis to examine the influence of vaccine efficacy (*ξ*) and the tendency for fast infection progression (*q*) on the intensity of medical mask distribution and case detection in TB control.

## 4. Sensitivity analysis

### 4.1. Global Sensitivity Analysis

This subsection is devoted to carrying out the global sensitivity analysis (GSA) of our Tb model. GSA is a process of investigating uncertainty analysis in a model output parameter given a model input factor over an entire range of interest. In addition, some advantages of using global sensitivity analyses are i) It considers all the input factors/parameters which are varied simultaneously while evaluating parameter sensitivity over the entire range of each input factor/time frame under investigation, (ii) It helps to identify model parameters that are more sensitive to infection threshold, which may be the infectious disease classes or control reproduction number as in our TB model, (iii) It also assesses the variability in model predictions, usually introduced by uncertainty in the parameter values. Knowing this information is relevant for policymaking in the management of the spread of both infectious and non-infectious diseases. To determine the variability in model parameters contained in control reproduction number, ℛ_*c*_, we use a combination of Latin Hypercube Sampling (LHS) and the Partial Rank Correlation Coefficients (PRCC) technique [48, 49]. Parameters with PRCC values above 0.5 or below -0.5 are the most significant or have strong correlations, which could be positive or negative respectively [48, 49]. This method looks at the relationship between ℛ _*c*_ and all its parameters. In this analysis, we use R software with 1000 simulations per run, and the resulting PRCC values indicate the effect of the parameters on the control reproduction number generated. These numerical results showing the PRCC for each parameter are shown as a Tornado plot in Figure 3 for Indonesia, India, Lesotho, and Angola, respectively.

**Figure 3:**
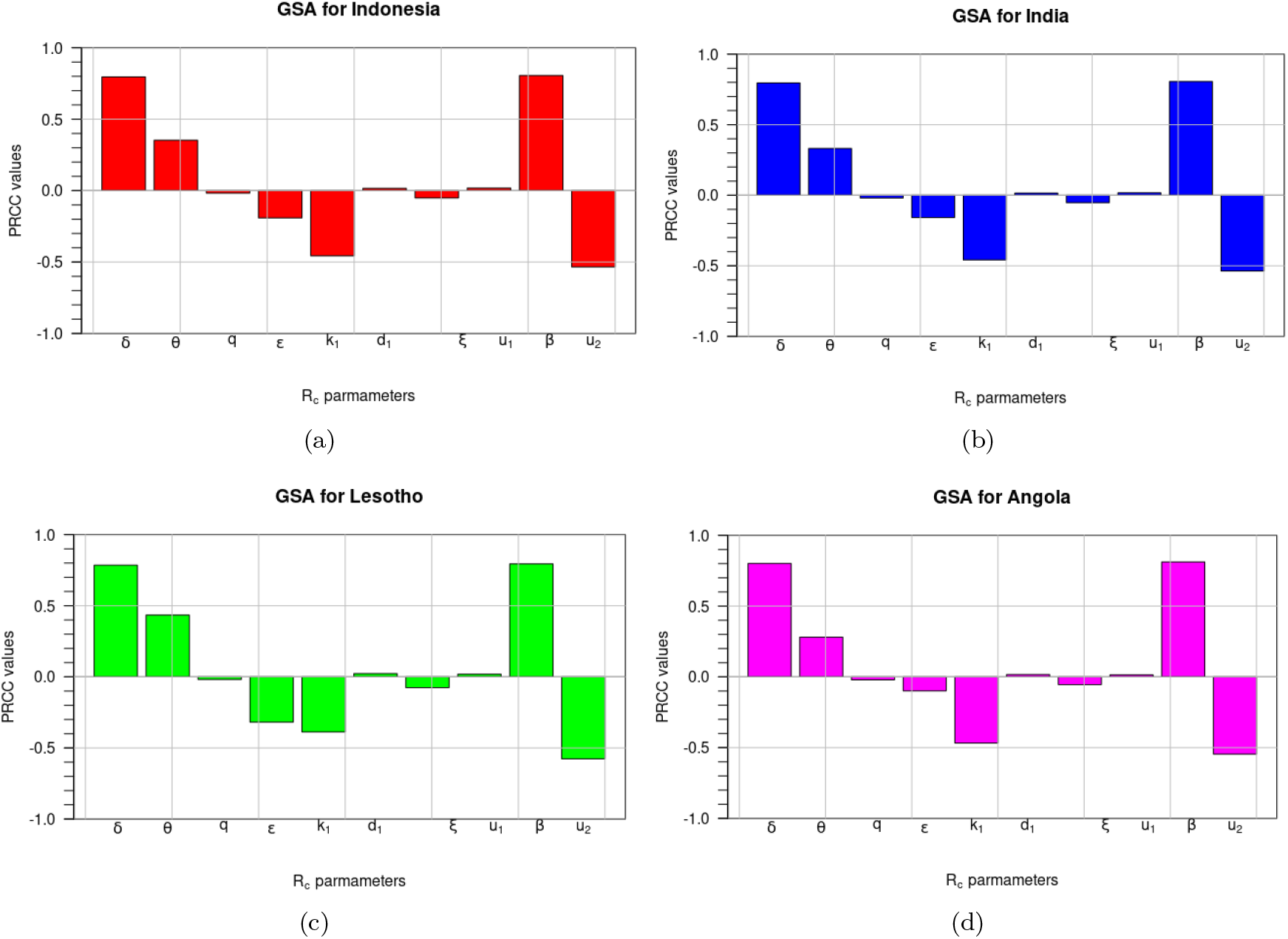
*Plots showing the global sensitivity analysis (GSA) on ℛ*_0_ *parameters excluding µ for:* (a) *Indonesia* (b) *India*, (c) *Lesotho* and (d) *Angola*.

In Figure 3(a), which represents the GSA results for Indonesia, the parameters *δ, β, u*_2_ and *θ* are the most sensitive. The infection rate *β* is positively correlated to ℛ _*c*_. This implies that it contributes to increasing the number of infectious individuals in the, which leads to more infected humans infected with Tuberculosis in Indonesia. Similar results apply to *δ* and *θ*. In contrast, the control parameter *u*_2_ has a negative correlation, which thus implies that effective mask usage reduces ℛ _*c*_ and, in turn, reduces the number of symptomatic infected TB individuals. A clear look at the results presented in Figures 3(b), 3(c) and 3(d) indicates that similar results are obtained for the TB-transmission path in India, Lesotho and Angola respectively. Many epidemic models in the literature have applied similar analyses to their, for instance, malaria [50, 51], Pneumonia [52], COVID-19 [53, 54], Listeriosis [55] and many others.

### 4.2. Effect of medical mask efficacy and fast-progression on the intensity of medical mask and case detection: A two-parameter sensitivity analysis

In this subsection, we conducted a two-parameter sensitivity analysis on ℛ _*c*_ with respect to the control variables *u*_1_ and *u*_2_, as well as the quality of medical mask parameter *ξ*. We considered two different values of *ξ* to represent the quality of a medical mask: *ξ* = 0.56 and *ξ* = 1. The value of *ξ* = 0.56 represents a condition that the medical mask offers 56% protection against the disease. On the other hand, if *ξ* = 1 represents a perfect quality of medical mask. All other parameter values are based on the best-fit parameter for Indonesia, which is shown in Table 1. The results are shown in Figure 4. Based on Figure 4 panel (a), an increase in the values of *u*_1_ and *u*_2_ increases the possibility of the value of ℛ _*c*_ becoming smaller than one. This is in line with the analysis as depicted in Figures F.7 and F.8.

**Figure 4:**
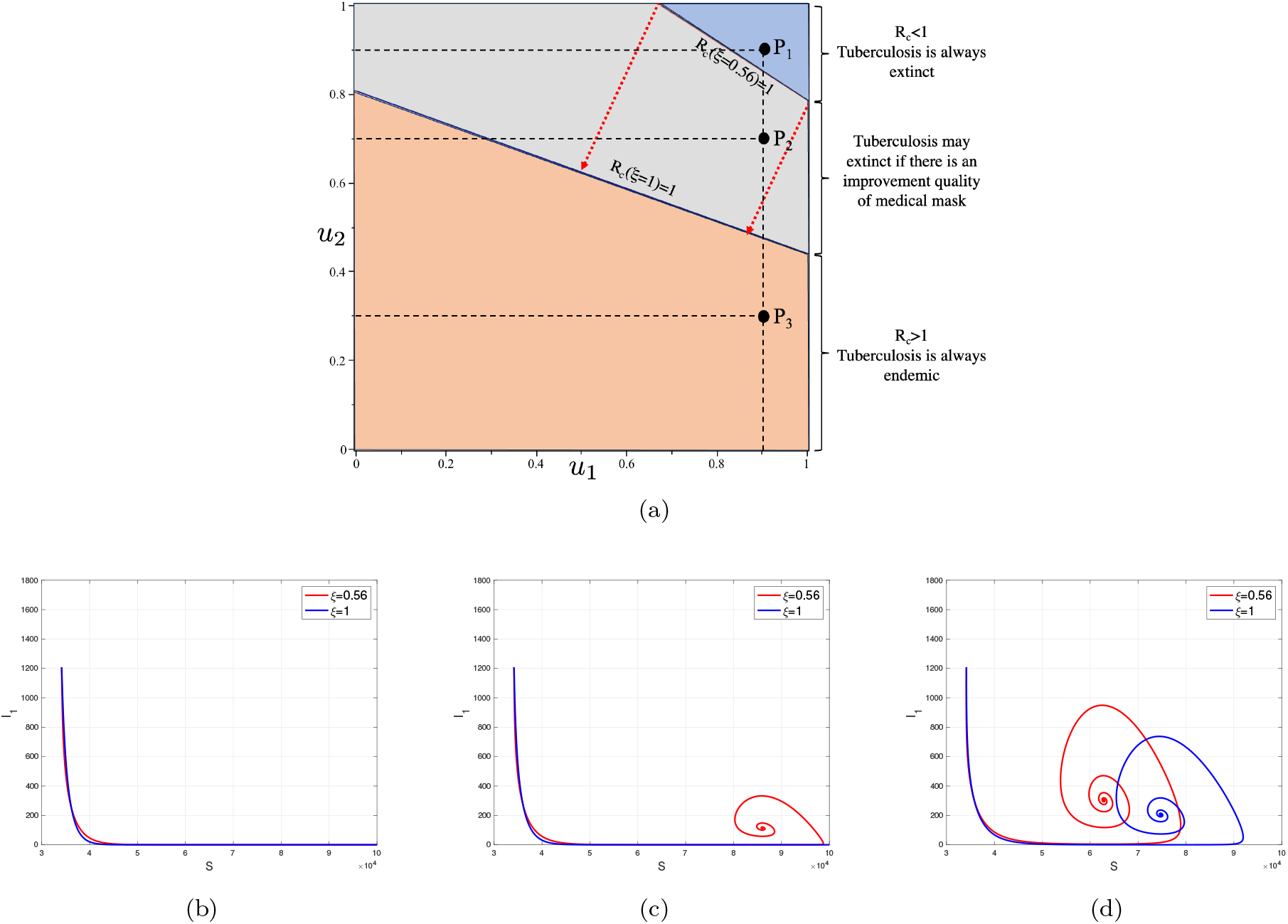
A two-parameter sensitivity analysis of ℛ _*c*_ respect to *u*_1_ and *u*_2_ under the impact of *ξ* given in panel (a). The orange region represents an ℛ _*c*_ domain that is always larger than one, while blue is always smaller than one, independent by the value of *ξ*. The gray region represents the area of ℛ _*c*_ that may change from greater than one to smaller than one as *ξ* increases. Panel (b), (c), and (d) show the dynamics of *S* and *I*_1_ for sample points *P*_1_, *P*_2_, and *P*_3_, respectively.

As mentioned before, the larger the values of *u*_1_ and *u*_2_, the greater the possibility that the value of ℛ _*c*_ becomes smaller than one. The orange-colored region represents the ℛ _*c*_ region, consistently exceeding one, while the blue-colored region signifies the ℛ _*c*_ region consistently remaining below one, despite a medical mask efficacy of only 56%. The gray area represents the range where ℛ _*c*_ could change from being greater than one (when *ξ* = 0.56) to being less than one (when *ξ* = 1). In other words, efforts related to the case detection rate (*u*_1_) or the proportion of individuals using medical masks (*u*_2_) can be minimized if the quality of the medical mask is improved. These findings also indicate that enhancing medical mask quality can indirectly contribute to reducing the required case detection rate, thereby controlling TB in the field.

To illustrate the influence of changes in *ξ* on the effectiveness of *u*_1_ and *u*_2_ regarding the variation of ℛ _*c*_ values, we selected three sample points, namely *P*_1_, *P*_2_, and *P*_3_, as shown in Figure 4 panel (b), (c) and (d). Point *P*_1_ is located in the blue region, where TB can be eradicated from the population irrespective of the quality of medical masks, whether it is 56% or 100%. Panel (b) illustrates that the dynamic of solutions consistently converges towards the TB-free equilibrium. On the contrary, Point *P*_3_ is situated in the orange area, where TB cannot be eliminated from the population regardless of the quality of the medical mask. Panel (d) demonstrates that the dynamic of *S* and *I*_1_ consistently tends toward the TB-endemic equilibrium. However, it is evident that a higher quality of medical mask results in a smaller endemic size of *I*_1_. Point *P*_2_ is located in the gray area, where TB elimination depends on the quality of the medical mask. If the medical mask efficacy is only 56%, then TB will persist in the population, as indicated by the red curve in panel (b) tending towards the TB-endemic equilibrium. Conversely, with a medical mask efficacy of 100%, TB can be eliminated from the population, as depicted by the blue curve in panel (b) converging towards the TB-free equilibrium.

Next, we analyze the sensitivity of *u*_1_ and *u*_2_ with respect to the value of ℛ _*c*_, using different values of the proportion of fast progression (*q*). A larger value of *q* indicates that more people proceed directly to active TB after their initial infection. Therefore, it is evident from Figure 5 panel (a) that a higher value of *q* will require more intense implementation of case detection and medical mask use to eliminate TB from the population.

**Figure 5:**
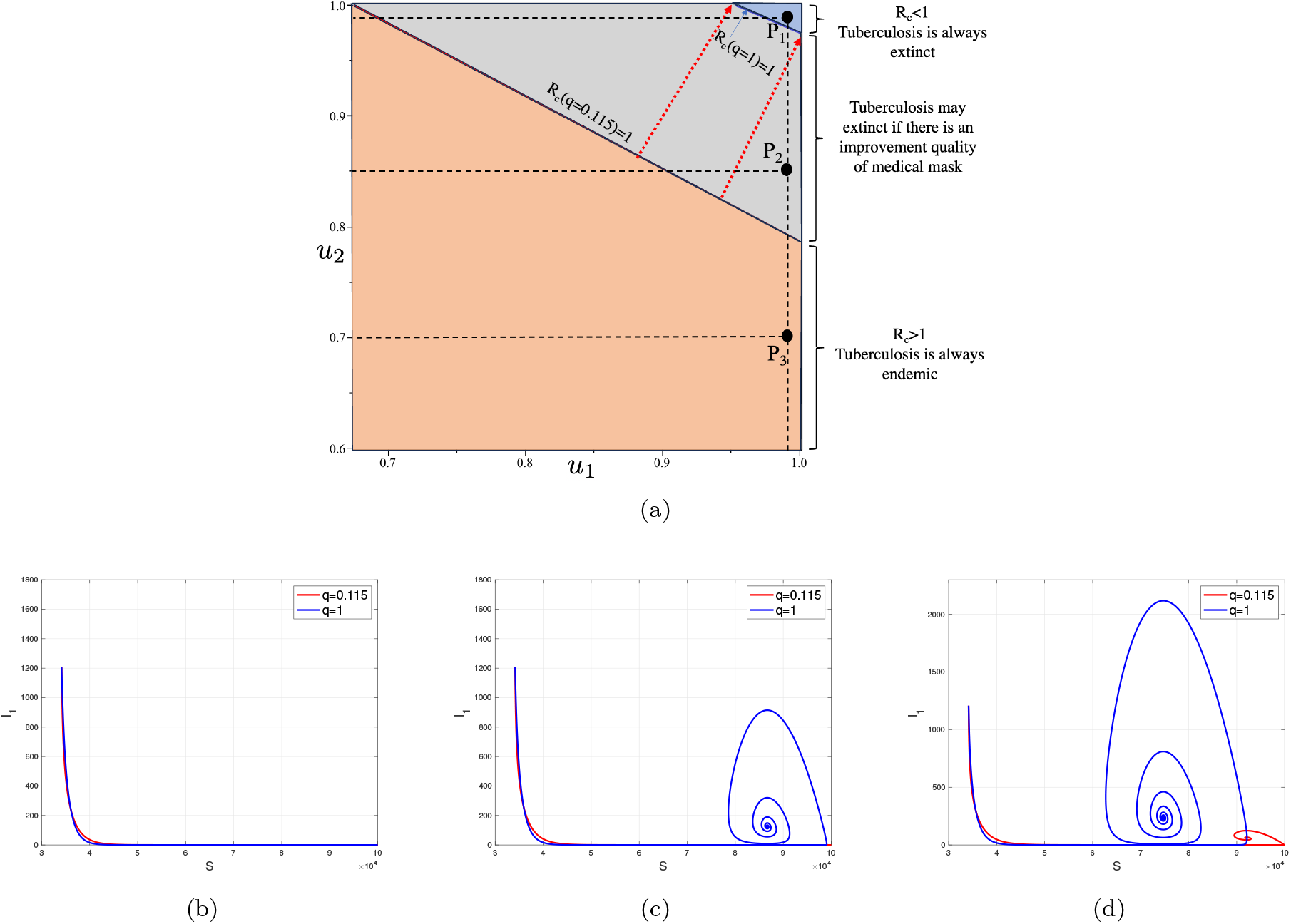
A two-parameter sensitivity analysis of ℛ_*c*_ with respect to *u*_1_ and *u*_2_ is presented under the influence of the parameter *q* (depicted in panel (a)). The orange region denotes the ℛ_*c*_ domain consistently exceeding one, while the blue region consistently remains below one, irrespective of the value of *q*. The gray region represents a zone where ℛ_*c*_ may either fall below or surpass one, contingent upon the value of *q*. Panels (b), (c), and (d) illustrate the dynamic behaviors of *S* and *I*_1_ for the sample points *P*_1_, *P*_2_, and *P*_3_” respectively.

We present an illustration of the dynamics of *S* and *I*_1_ based on various combinations of *u*_1_, *u*_2_, and *q*. In the case of the combination in *P*_1_ (as shown in Figure 5 panel (b)), both the dynamics of *S* and *I*_1_ tend towards the TB-free equilibrium point. A smaller value of *q* accelerates the convergence of both *S* and *I*_1_ towards the TB-free equilibrium point. Similarly, in Figure 5 panel (d), both dynamics tend towards the TB-endemic equilibrium point, as ℛ_*c*_ *>* 1 at *P*_3_. However, it is important to note that *P*_2_ (shown in Figure 5 panel (b)) does not always result in ℛ_*c*_ *<* 1. For instance, when *q* = 0.115, the combination of *u*_1_ and *u*_2_ at *P*_2_ yields ℛ _*c*_ *<* 1, leading to dynamics that approach the TB-free equilibrium point (see the red curve). Conversely, when *q* = 1, the combination of *u*_1_ and *u*_2_ at *P*_2_ results in ℛ_*c*_ *>* 1, causing the dynamics of *S* and *I*_1_ tend towards the TB-endemic equilibrium point (see the blue curve).

## 5. Optimal Control Model of Case Detection and Medical Mask

### 5.1. Optimal Control Characterization

As mentioned in the previous analysis, it is clear that a more substantial intervention in case detection and medical mask usage will significantly reduce both the size of the control reproduction number and the size of the infected compartment in the TB-endemic equilibrium. However, the more extensive the intervention, the higher the cost. Therefore, the implementation of case detection and medical mask usage should adapt to the condition of the infected compartment over time.

This section treats the control intervention as time-dependent variables, denoted as *u*_1_ = *u*_1_(*t*) and *u*_2_ = *u*_2_(*t*). Consequently, the model in (1) now reads as:

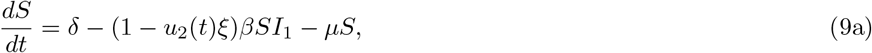

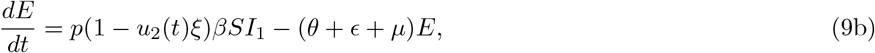

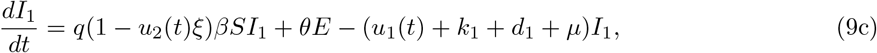

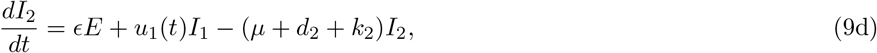

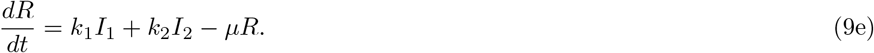

We aim to minimize the cost function as follows:

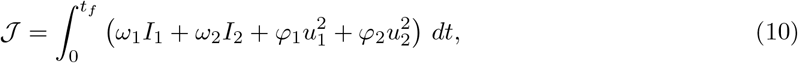

where *ω*_1_, *ω*_2_ are the weight parameters for infected compartment, while *φ*_1_ and *φ*_2_ are the weight costs for the control variables. Each component on 𝒥 can be described as follows:

- The cost due to all interventions, except the use of medical masks and case detection, in controlling the number of infected individuals *I*_1_ and *I*_2_ is described by 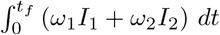.
- The cost, due to the intensity of intervention implemented, is given by 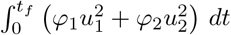.

This optimal control construction aims to seek an optimal trajectory for 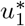 and 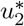 to minimize the cost function 𝒥. Mathematically, it is described by the following equation

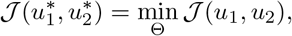

where Θ = {(*u*_1_, *u*_2_)|*u*_*i*_ is Lebesgue measurable function,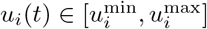} is the set of admissible control. By applying the Pontryagin’s Maximum Principle, we define the Hamiltonian function as follows:

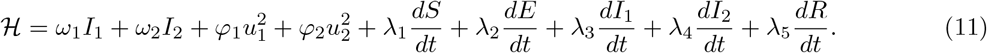

With this Hamiltonian function, we have the following Theorem 6.

#### Theorem 6.

*Let the solution of the optimal control problem are* 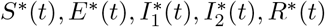 *with it* 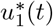 *and* 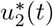. *Then there exist an adjoint variables λ*_*i*_ *for i* = 1, 2, 3, 4, 5 *which satisfies the following system:*

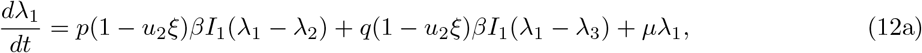

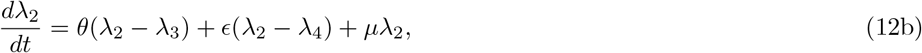

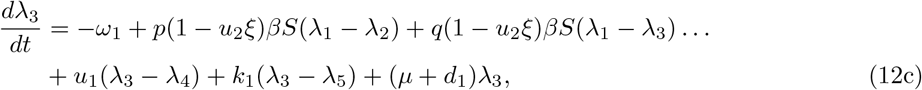

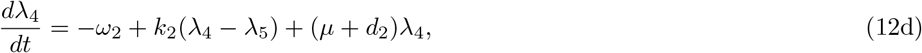

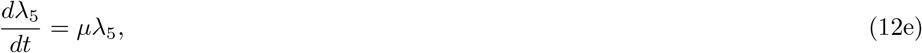

*with their transversality conditions λ*_*i*_(*t*_*f*_) = 0 *for i* = 1, 2, 3, 4, 5. *The optimal trajectory on its admissible set is given by*

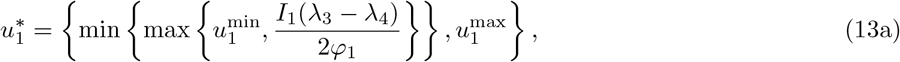

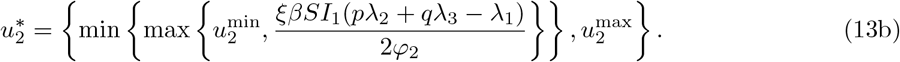

For the derivation of Theorem 6, readers may refer to Appendix G for the complete proof.

### 5.2. Numerical Experiments of the Optimal Control Problem

To solve the optimal control problem, we employed the forward-backward sweep method. Model (9) was solved using forward sweep, a set of initial guesses for the control variables. Then, the adjoint system in equation (12) was solved backward sweep using the initial guess and the solution obtained from the previous step. Using the results, we computed the control variables as described in equation (13). All the steps were repeated until the convergence criteria were achieved. Further details and practical examples of the method can be found in [8, 43].

The numerical experiments in this section encompassed two distinct scenarios. The first scenario involved forecasting the incidence of cases in Indonesia, India, Lesotho, and Angola for 30 years onward up to 2050, with the optimal control variables incorporated into the model from 2021 to 2050. The second experiment focused on simulating various scenarios for control implementation, including case detection only, using medical masks only, and combining both interventions.

#### 5.2.1. Forecast of TB case incidence with time-dependent intervention

The numerical experiments conducted in this section utilized the best-fit parameters in Table 1. We forecasted the case incidence for each country until 2050 by implementing both case detection and the use of medical masks. The results for Indonesia are presented in Figure 6. These forecasting results for India, Lesotho, and Angola can be found in Figures H.10, H.11, and H.12 in Appendix H.

**Figure 6:**
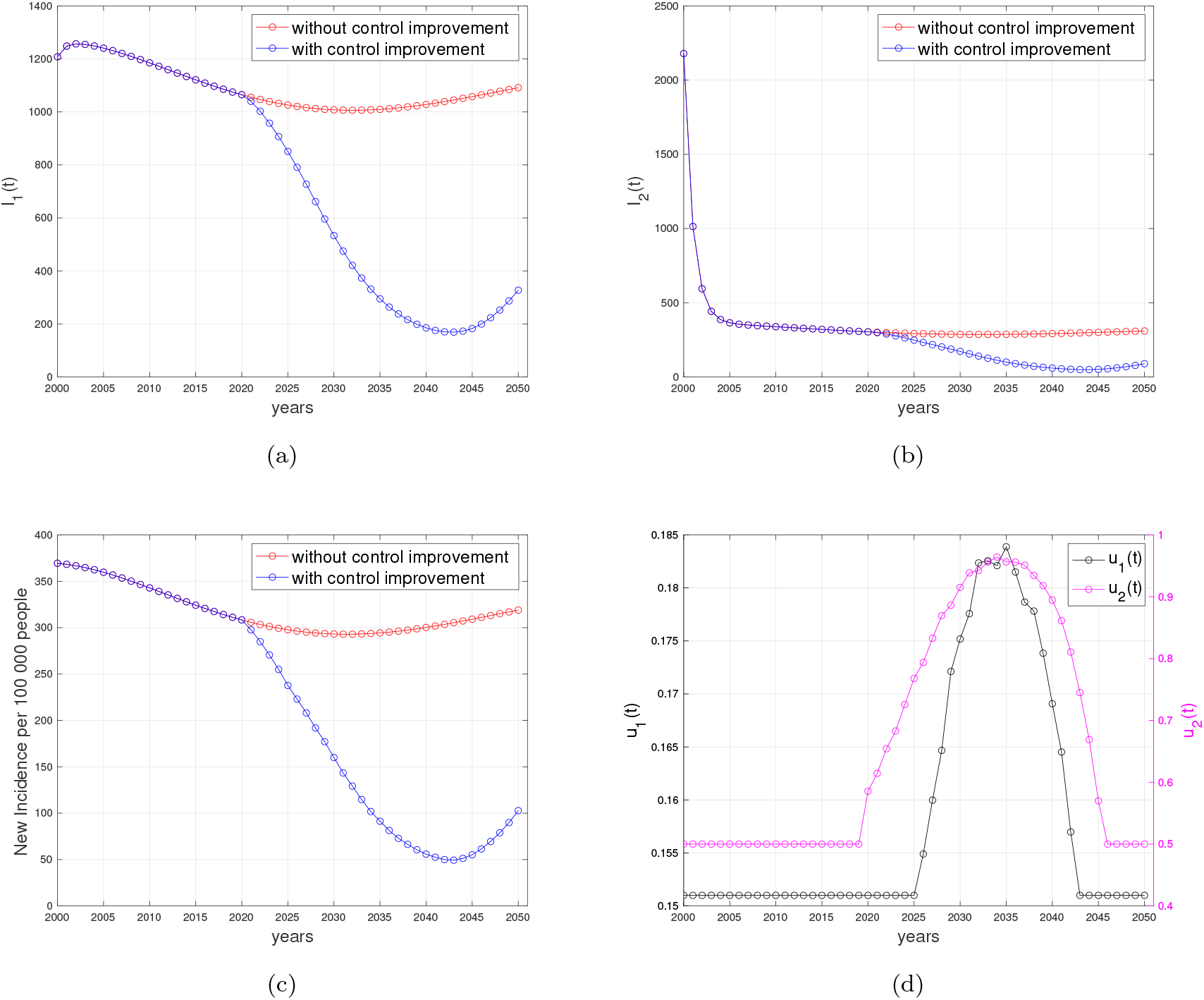
Forecasting and optimal control results for Indonesian data. Panels (a) and (b) represent the dynamic of *I*_1_ and *I*_2_, respectively. Panel (c) represents the case incidence per 100,000 people without and with control improvement, while panel (d) shows the dynamic of control parameters.

Based on the results in Figures 6 (a) and (b), it is evident that the number of infected individuals continued to decrease even more significantly when control interventions were improved from 2021 until 2050, as control dynamics depicted in Figure 6 (d). The control interventions from 2000 to 2020 used the data shown in Table 1. This trend corresponds directly to the prediction of case incidence per 100,000 population in panel (c), where it decreases significantly when control interventions improve but begins to rise when control interventions are reduced. It is worth highlighting that the substantial reduction shown in Figure 6 (c) starting from the year 2021 was due to active case finding (corresponding to *u*_1_) within the population. Mathematically, this sharp reduction was because of rapid transitions from the *I*_1_ compartment to the *I*_2_ compartment following the medical mask intervention. Without this *u*_1_, the number of detected cases was mainly contributed by the *E* compartment alone. Case detection should be improved from 2021, and it should start to decrease from 2035 onward to minimize the intervention cost. Furthermore, we can see clearly that when the control interventions were gradually removed, then the number of cases began to rise again, highlighting their importance in controlling the outbreak. A similar pattern was observed in numerical simulations results for India, Lesotho and Angola, where the number of infected individuals decreased gradually with and without the control interventions, as shown in Figure H.10, H.11, and H.12 in Appendix. Nevertheless, implementing control interventions speeds up the eradication process of the infected cases, compared to the case of no control intervention.

There are Several important notes from numerical experiments for India, Lesotho and Angola. For simulation results using data from India, it is evident that interventions must be significantly enhanced since 2021 and start to decline in the year 2040. In an extreme scenario, the proportion of the population required to use medical masks reaches 100% from 2034 to 2040 and then begins to decrease significantly to 50%. Regarding data from Lesotho, no significant increase was observed in case detection interventions; it remained constant. Conversely, the intervention for medical mask usage needs to be significantly increased starting in 2021 and then starting to decline in 2033. There is apparent no significant decrease in the new incidence rate in Lesotho with such control dynamics. Analyzing data from Angola, it is evident that the number of people using medical masks must have significantly increased since 2021, even reaching 100% from that year onwards. This intervention starts to decline in the year 2041. On the other hand, the intervention for case detection must also be increased since 2021. However, the difference lies in the fact that the intervention for case detection begins to decline in 2033 to offset the high intervention of medical mask usage, which undoubtedly incurs non-trivial costs.

#### 5.2.2. Assessing the effectiveness of various combinations of control strategies

The model incorporates two different control strategies: case detection and medical mask usage. As previously explained, case detection and medical mask interventions have different focuses. The case detection intervention aims to actively identify active TB cases in the field and provide them with appropriate treatment. On the other hand, the medical mask intervention is more oriented towards preventing the spread of TB by encouraging infected individuals to protect the population by using medical masks. In essence, case detection is a mitigation intervention, while medical mask use is a preventative measure.

The effectiveness of various combinations of these control strategies was investigated in this section. In particular, the following three scenarios were considered in our numerical simulations:

1. Implementation of both case detection and medical mask usage i.e *u*_1_(*t*) ∈ [0.151, 1] and *u*_2_(*t*)[0.5, 1].
2. Implementation of case detection only i.e *u*_1_(*t*) ∈ [0.151, 1] and *u*_2_(*t*) = 0.5.
3. Implementation of medical mask usage only i.e *u*_1_(*t*) = 0.151 and *u*_2_(*t*) ∈ [0.5, 1].

The lower bounds of each control parameter are derived from the best-fit parameters outlined in Table 1, whereas the upper bounds are set equal to one. All numerical results are given in Appendix Appendix I. In each panel depicting the dynamics of every compartment from (a) to (e), we assess three distinct scenarios: firstly, when no controls were implemented (*u*_1_ = *u*_2_ = 0); secondly, when interventions regarding case detection and the utilization of medical masks remained constant until the year 2051; and thirdly, when improvements were made in case detection and/or medical mask utilization.

##### Scenario 1: improvement in case detection and medical mask use

The results of the first scenario are presented in Figure I.13, where both case detection and medical mask interventions were simultaneously implemented. Figures I.13 (a)-(e) depict the dynamics of compartments *S, E, I*_1_, *I*_2_, and *R*, respectively. It is evident that by improving both case detection and medical mask use interventions, as shown in Figure I.13 (g) and (h), the number of healthy individuals can be significantly increased, and the number of infected individuals can be significantly reduced. As a result, it can be observed that the number of detected infected individuals steadily decreased in proportion to the decrease in the number of infected individuals (see Figure I.13 (f)). However, as the intervention diminishes from 2036 onwards, the incidence of new cases rises, particularly evident from 2046. Unlike the intervention of medical masks, which requires improvement since 2021, the intervention of case detection may remain constant at *u*_1_ = 0.151 from 2021 until 2026, subsequently increasing until 2036. Under this scenario, the total number of averted infections reaches 1.226 *×* 10^5^.

##### Scenario 2: improvement in case detection only

In this scenario, we conducted numerical simulations to understand the effects of case detection as a single intervention that we need to improve in controlling TB transmission in Indonesia. In contrast, the intervention of medical mask use remains constant at *u*_2_ = 0.5. The results are presented in Figure I.14. The case detection rate in this scenario is illustrated in Figure I.14 (g), which is significantly higher compared to the previous scenario where case detection intervention was accompanied by medical mask usage in TB mitigation efforts (see Figure I.13 (g)). Despite the intensive implementation of case detection intervention, the reduction in the number of infected individuals was not as effective as in the previous scenario (see Figure I.14 (a) to (e)). This scenario’s total number of infections averted was 8.9 *×* 10^4^.

##### Scenario 3: improvement in medical mask use only

For the last scenario, we examined the impact of the improvement on medical mask use in preventing the spread of TB while keeping the case detection constant at *u*_1_ = 0.151. The results of the numerical simulation are shown in Figure I.15. Since the use of medical masks was the sole intervention in this scenario, the intervention intensity was higher compared to the first scenario, as indicated in Figure I.15 (h). We can see that the intervention of medical masks alone reduced the total number of infected cases better than the intervention of case detection alone. This is supported by the fact that this scenario’s total number of infections averted was 1.295 *×* 10^4^. This was smaller than the first two scenarios.

##### Cost-Effectiveness Analysis

Based on the numerical simulations conducted for all three scenarios mentioned above, implementing both interventions (scenario 3) yields the most significant results in the number of infections prevented, but it is only slightly different from scenario 1, where both interventions improved. However, this may come at a higher intervention cost. Therefore, an analysis was needed to assess the effectiveness of these strategies relative to the costs incurred. To do so, we used the Average Cost-Effectiveness Ratio (ACER) to compare the effectiveness of our three scenarios. The formula to calculate ACER is given by

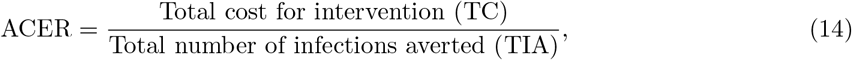

The formula for ACER is given by (14), where TC is the total cost of interventions *u*_1_ and *u*_2_, while TIA is the total number of infections averted from compartments *E, I*_1_, and *I*_2_. A smaller value of ACER represents a more cost-effective strategy.

Table 2 shows that ACER for scenario 1 was the smallest, followed by scenarios 2 and 3. Our results demonstrated that combining both interventions provided better cost-effectiveness than the other two scenarios.

**Table 2:** The cost-effectiveness analysis (ACER) for all three scenarios.

| Scenario | TC | TIA | ACER |
| --- | --- | --- | --- |
| 1 | $1.281 \times 10^4$ | $1.226 \times 10^5$ | 0.1044 |
| 2 | $1.878 \times 10^4$ | $8.9 \times 10^4$ | 0.211 |
| 3 | $1.295 \times 10^4$ | $1.227 \times 10^5$ | 0.1055 |

The next cost-effectiveness analysis is the IAR (Infection Averted Ratio) analysis. This analysis assessed the effectiveness of interventions to prevent the spread of infectious diseases. It quantifies the impact of an intervention by comparing the number of infections averted due to the intervention to the total number of recovered individuals due to the implementation of control programs. Hence, the formula to calculate IAR is given by:

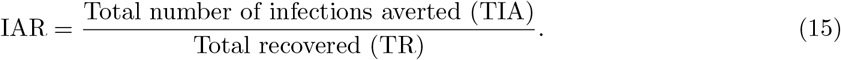

A higher IAR value indicates that the intervention is more efficient in preventing infections relative to the recovered individual. The IAR analysis helps decision-makers prioritize interventions and allocate resources effectively to maximize the impact of disease prevention efforts. Using this formula, we calculate the IAR for each scenario and yield that the IAR for scenario 1 is 0.199, scenario 2 is 0.1228, and scenario 3 is 0.2. Hence, we find that the intervention of medical mask use alone is the best strategy using the IAR indicator, followed by the combination of medical mask and case detection, and lastly, medical mask use alone.

From the above cost-effectiveness analysis and the global sensitivity analysis, we can see that implementing medical masks is more successful in reducing the number of infected individuals than in case detection. However, we need to be careful that there are several reasons why medical masks may be considered a better strategy for TB control compared to case detection in certain situations. First, a medical mask is a preventive measure by reducing the probability of successful TB transmission. Medical mask use can help prevent the spread of TB in crowded or poorly ventilated settings, such as healthcare facilities, prisons, and shelters, where close contact between individuals increases the risk of TB transmission. In contrast, case detection primarily targets identifying and treating individuals already infected with TB, which may not effectively prevent transmission from undetected cases. Second, medical masks are relatively more universal in their applications and more efficient in the cost of implementation. In resource-limited settings where healthcare infrastructure is limited, medical masks may offer a cost-effective approach to TB control. It is important to note that while medical masks can play a valuable role in TB control, they are not a standalone solution and should be integrated into comprehensive TB control programs alongside other interventions, including case detection, treatment, infection control measures, and public health education.

## 6. Discussion and Conclusion

A TB model that includes the effects of medical mask use and active case finding is proposed in this article. This nonlinear system of ordinary differential equations consists of five variables representing various classes of the human population and 14 parameters. The analysis of the mathematical model concerning the threshold reproduction number, ℛ _*c*_, demonstrates that implementing medical mask usage and active case finding can potentially mitigate the spread of TB effectively. From equilibrium analysis, we find that our model always exhibits a transcritical bifurcation at ℛ _*c*_ = 1. This finding suggests that the persistence of TB occurs when the reproductive number, ℛ _*c*_ *>* 1, while TB will become extinct if ℛ_*c*_ *<* 1 (see Figure F.7, F.8, and F.9).

Our model is parameterized using yearly data on TB cases per 100000 population from four different countries: Indonesia, India, Lesotho, and Angola (see section 2.2). This parameterization revealed that the reproduction numbers for all four countries consistently exceeded one, suggesting the potential for sustained TB endemicity. Nevertheless, our model’s forecasting results indicated a declining trend in tuberculosis cases within these countries in the coming years (refer to Figures 6, H.10, H.11, and H.12).

Numerical experiments have been carried out to provide evidence that medical masks and proactive case identification can significantly enhance the potential of eliminating TB from the population. As parameter *ξ* indicated within our model, a better quality of medical masks also contributed to the reduction of ℛ _*c*_. In addition, as the probability of fast disease progression, *q* increased, a more intense utilization of medical masks and proactive efforts by the government to conduct active case finding became imperative for the effective eradication of TB. Our sensitivity analysis showed that undetected and detected active TB dynamics were highly responsive to active case findings. This sensitivity was most pronounced at the outset of the intervention and gradually diminished over time.

Using this parameterization, we further developed our model into an optimal control model to analyze the effectiveness of case detection and usage of medical masks as a function of time in controlling TB. From the results obtained, it is evident that the combination of both case detection and the usage of medical masks proved to be a cost-effective and effective strategy for mitigating the spread of TB, notably in reducing the incidence of infection. These findings can provide valuable insights into the complexity of TB transmission in these four countries. The differing focus of interventions can be considered in the future and adapted to the on-ground conditions.

Our research shows a big potential for medical masks to be used as a non-pharmaceutical intervention for TB eradication programs (even with a small efficacy of only 56% [34]). Implementing medical masks as a tuberculosis (TB) intervention over a 50-year eradication program presents several challenges and considerations. While medical masks may offer some degree of protection against TB transmission, their long-term feasibility and effectiveness in such a program would depend on various factors such as acceptance and compliance, access and affordability, and cultural and social factors. Implementing medical masks for such a long period may require significant efforts to promote awareness, education, and behavior change. Ensuring consistent compliance with mask-wearing guidelines over many years could be challenging, especially in regions with low TB prevalence or where perceptions of risk fluctuate. Furthermore, governments and health authorities would need to invest in infrastructure, supply chains, and subsidies to make medical masks accessible to all socio-economic groups over the long term. In summary, while implementing medical masks as a TB intervention over a 50-year eradication program is theoretically feasible, it would require comprehensive planning, sustained investment, community engagement, and integration with broader TB control strategies. Although it is a challenging effort to implement this on a big scale, like on a country scale, the implementation of medical mask usage can begin at the grassroots level, starting with the smallest social circles such as households with TB-infected individuals, hospitals [56], and similar environments.

As mentioned earlier, the challenge in TB control globally persists due to several factors. One major issue is the emergence of drug-resistant TB strains, such as multidrug-resistant TB (MDR-TB) and extensively drug-resistant TB (XDR-TB), which are harder and costlier to treat. Additionally, TB often affects marginalized and vulnerable populations, making access to healthcare and proper diagnosis a challenge. Addressing these challenges requires a multi-pronged approach, including augmented financial resources, enhanced diagnostic capabilities, and strengthened healthcare systems to ensure equitable access to TB care. Hence, it is important to continue our efforts to refine the model by integrating the factors above into our new model in the future.

## Data Availability

T. W. Bank, Incidence of tuberculosis (per 100,000 people), https://api.worldbank.org/v2/en/
indicator/SH.TBS.INCD?downloadformat=excel (2023)

https://api.worldbank.org/v2/en/indicator/SH.TBS.INCD?downloadformat=excel

## Acknowledgement

The authors thank all reviewers for their valuable comments that helped to improve this manuscript. Universitas Indonesia funds this research through the PUTI research grant scheme 2023 (NKB-469/UN2.RST/HKP.05.00/2023).

## Appendix A. Proof of invariant region Ω

The invariant region of the proposed model can be shown below, following the approach in [57, 58]. The solutions with non-negative initial conditions remain in a neighborhood of the closed positive hyperspace, 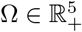, for *t* ≥ 0. That is, 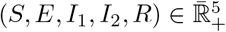, for

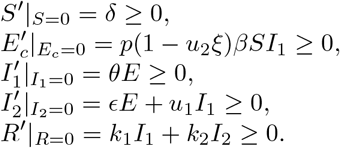

It can be seen that the gradient on the boundary of 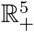 is always positive. Hence, the solution will always non-negative for all *t >* 0. Summing all right hand side of equations in (1), we have:

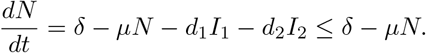

Hence, using integrating factor method, the solution of *N* satisfying:

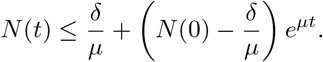

It can be seen that if 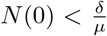, then *N* (*t*) will monotonically increasing and tends to 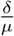. If 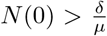, then *N* (*t*) will monotonically decreasing and tends to 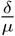. If 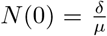, then *N* (*t*) always stays inside of Ω. Therefore, all feasible solution of the model (1) enter the region Ω implying that the region is an attracting set. Hence the proof is complete.

## Appendix B. Proof of Theorem (1): Derivation of the control reproduction number

Using the Next Generation Matrix (NGM) method [59], here are steps to find ℛ _*c*_.

1. Create the Jacobian matrix **J** of the infected compartment by linearization around the DFE point as follows.

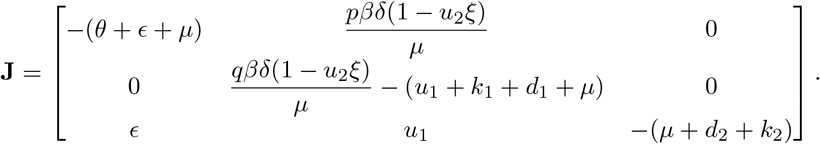
2. Decompose the Jacobian matrix as **J** = **T** + Σ with **T** is the transmission matrix and Σ is the transition matrix as follows.

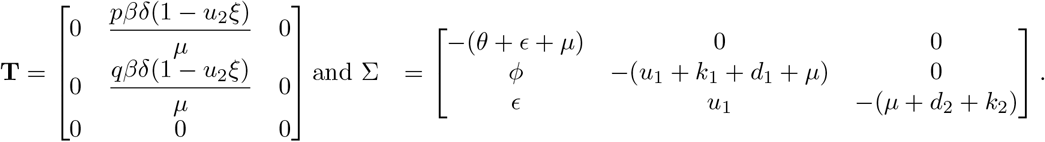
3. As we can see, the **T** matrix consists of one row that is entirely zeros, then we must construct the **E** matrix, which columns are unit vectors that correspond with a non-zero row of **T**.

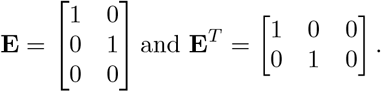

Use one of NGM’s formulas that is denoted by the **K** matrix as follows.

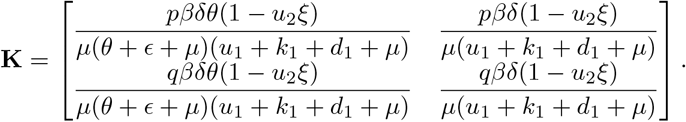

From the **K** matrix, we get the determinant of **K** is zero. Therefore, we use another one of NGM’s formulas involving a small domain. Hence, ℛ _*c*_ is given by

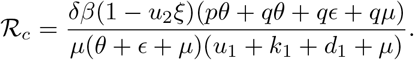

## Appendix C. Proof of Theorem (2): Local stability criteria of *E*_1_

We use Van den Driessche and Watmough’s approach [45] to analyze the local stability of DFE. First, define **x** = (*x*_1_, *x*_2_, *x*_3_, *x*_4_, *x*_5_)^*T*^ with *x*_1_, *x*_2_, *x*_3_, *x*_4_, and *x*_5_ is the number of individuals in each compartment *E, I*_1_, *I*_2_, *S* and *R* respectively. Then, define **X**_*s*_ as the set of DFE populations.

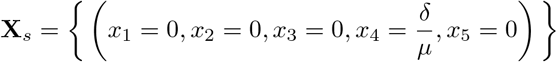

Define *f*_*i*_(*x*) for *i* = 1, 2, .., 5 which represents 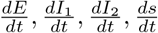, and 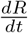. Note that the initial condition for each population must have a non-negative value following this condition.

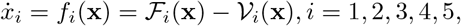

Then, we have

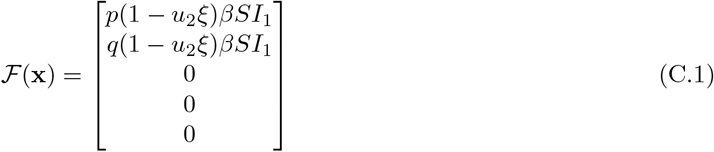

and

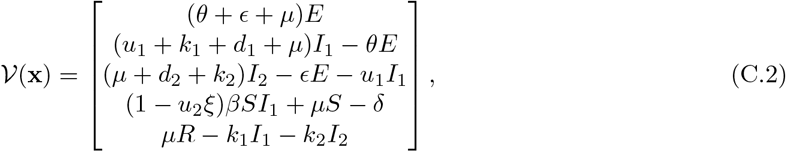

where 𝒱 (**x**) = 𝒱 ^−^(**x**) − 𝒱 ^+^(**x**) are given by

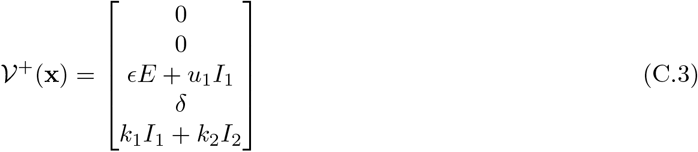

and

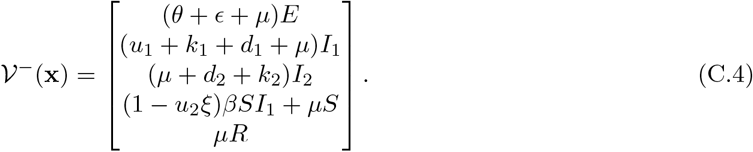

After that, analyze whether DFE is locally asymptotically stable when ℛ_*c*_ *<* 1 using five axioms from Van den Driessche and Watmough.

1. If *x*_*i*_ ≥ 0, then 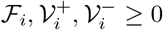 for *i* = 1, 2, …, 5. All parameters and variables have non-negative input. Based on that, the first axiom is satisfied.
2. If *x*_*i*_ = 0, then 𝒱 ^−^ = 0 for *i* = 1, 2, 3. By substituting *x*_*i*_ = 0 for *i* = 1, 2, 3 to equation (7) we will get 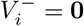. Hence, the second axiom is satisfied.
3. ℱ_*i*_ = 0 if *i >* 3. For *i* = 4, 5, as we can see from equation (4) that ℱ_4_ = ℱ_5_ = 0, then the third axiom is satisfied.
4. If *x*_*i*_ ∈ **X**_*s*_, then ℱ_*i*_ = **0** and 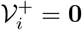 for *i* = 1, 2, 3. Substitute *x*_*i*_ = 0 to equation (4) and (6), then we will get ℱ_*i*_ = **0** and 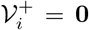 for *i* = 1, 2, 3. Hence, the fourth axiom is satisfied.
5. If ℱ_*i*_ = **0**, then all eigenvalues of *D****f*** (*x*_0_) have a negative real part. By substituting ℱ_*i*_(**x) = 0** to *f*_*i*_(*x*), we obtain

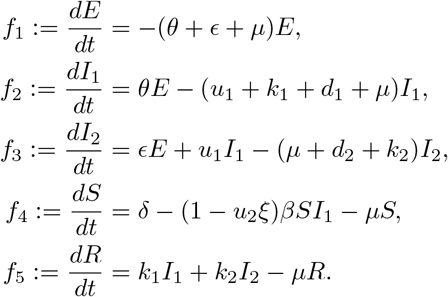

Then, we obtain the Jacobian matrix of *f*_*i*_(*x*) that is evaluated at DFE as follows.

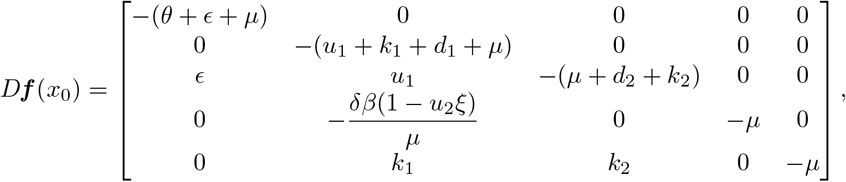

with eigenvalues of matrix above are −(*θ* + *ϵ* + *µ*), −(*u*_1_ + *k*_1_ + *d*_1_ + *µ*), −(*µ* + *d*_2_ + *k*_2_), −*µ*, and −*µ*. Note that all parameters have non-negative values, it makes all eigenvalues have a negative real part. Then, we can conclude that the last axiom is satisfied too.

According to these results, all axioms are satisfied. Furthermore, we can guarantee in the second theorem that the disease-free equilibrium of system (1) is locally asymptotically stable.

## Appendix D. Proof of Theorem 3: Global Stability of the Disease-free equilibrium

To prove that the DFE is GAS whenever ℛ_0_ *<* 1, we have to verify the conditions 𝒞_1_ to 𝒞_3_. Using the result in [45], we obtain that the DFE ℰ_1_ is LAS when ℛ_0_ *<* 1, so the condition 𝒞_1_ is verified. Next, we re-write the model system (1) in the form given in (5) as

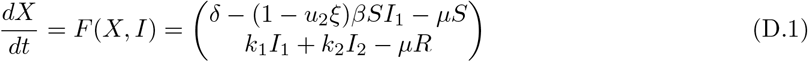

and

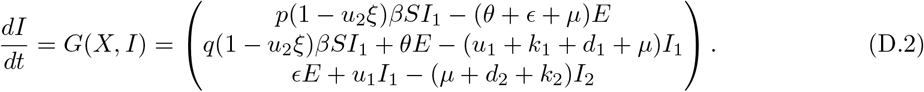

We have that

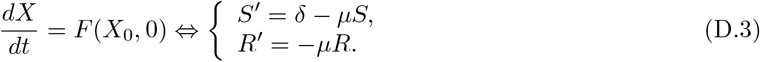

Equation (D.3) has a unique equilibrium point 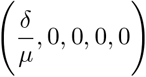 which is globally asymptotically stable.

Hence, the condition 𝒞_2_ is satisfied. Linearizing equations (D.1) and (D.2) yields the Metzler Matrix (𝒜 = 𝒟_*Z*_*𝒢* (ℰ_1_)) given as

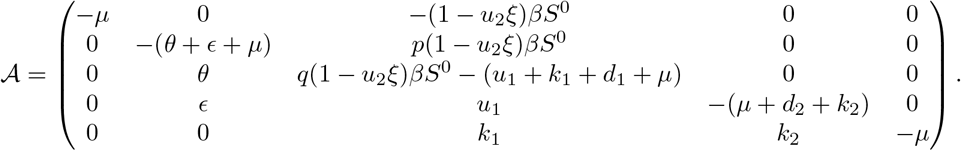

Computing 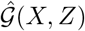 and after some algebraic manipulation, we have that

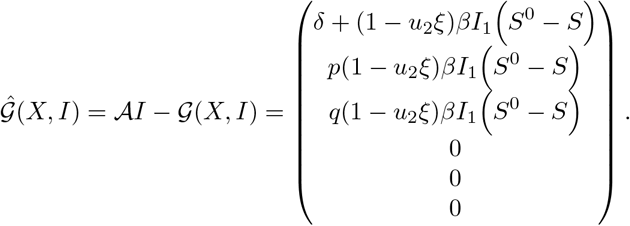

Thus,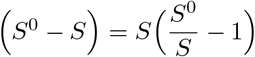 is positive due to our assumption, and hence 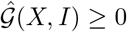. The condition 𝒞_3_ is also satisfied. We can conclude that our model is globally asymptotically stable.

## Appendix E. Proof of Theorem (5): Non-existence of backward bifurcation

We use Castillo-Cahvez and Song theorem [47] to analyze the stability of endemic equilibrium from the system (1). First, suppose each compartment is as follows.

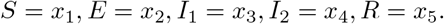

Then, redefine the system as,

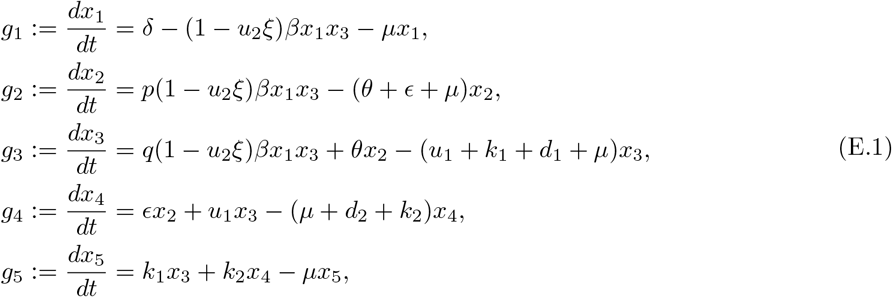

Choose *β* as the bifurcation parameter, then setting ℛ_*c*_ = 1 and we obtain,

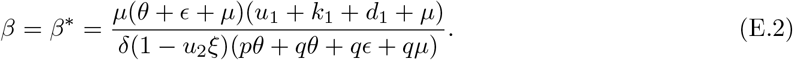

Linearization system of equation (E.1) at DFE with *β* = *β*^∗^ as follows.

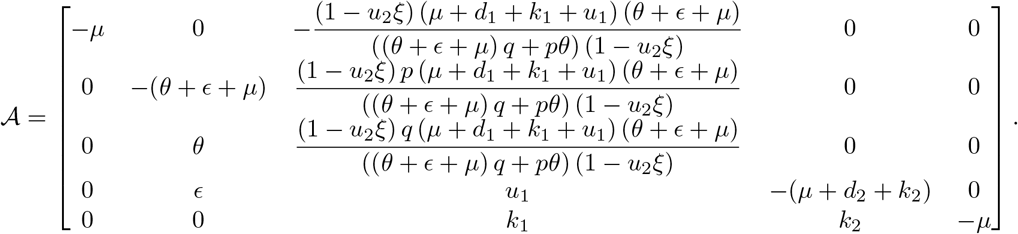

From the Jacobian matrix, we get a simple zero eigenvalue (with all other eigenvalues having negative real parts). Hence, we can continue our step by computing the right eigenvector **w** of matrix 𝒜 that correspondents to *λ* = 0, which satisfied 𝒜**w** = *λ***w**. After that, we have

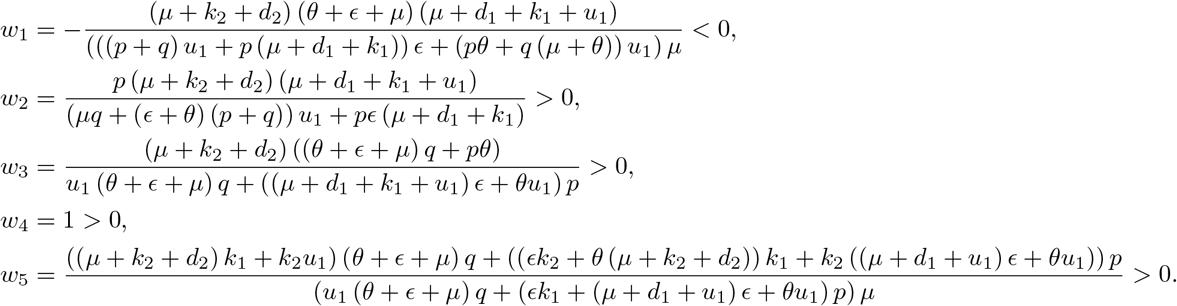

Next, compute the left eigenvector **v** of matrix 𝒜 that correspondents to *λ* = 0, which satisfied **v𝒜** = **v***λ*, then we have

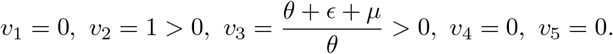

The next step is calculating the value of *a* and *b* as follows.

1. As we know that *v*_1_ = *v*_4_ = *v*_5_ = 0, then *g*_1_, *g*_4_, and *g*_5_ are not considered in calculation. Therefore, we just consider *g*_2_ and *g*_3_ in this calculation. Hence, we have the bifurcation coefficient *a* given by.

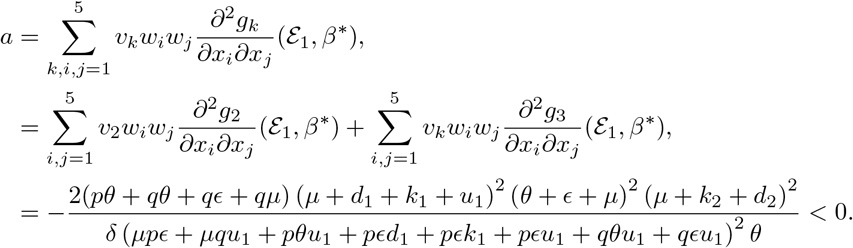
2. Note that *v*_1_ = *v*_4_ = *v*_5_ = 0, then *g*_1_, *g*_4_, and *g*_5_ are not considered in this calculation. Hence, the bifurcation coefficient *b* is given by.

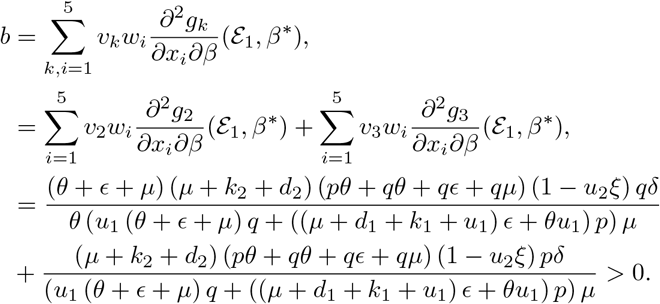 Because *a* is negative and *b* is positive, then by Catillo-Chavez and Song theorem the system (1) exhibits the phenomena of forward bifurcation at ℛ_*c*_ = 1.

## Appendix F. Numerical experiments on bifurcation diagram and autonomous simulation

Here in this section, we performed the sensitivity analysis on the bifurcation diagram of system (1) with respect to control parameters *u*_1_ and *u*_2_ and the infection rate *β*. The results are displayed in Figures F.7-F.9. The parameter values employed for numerical simulations in this section correspond to the optimized values for Indonesia, detailed in Table 1, unless explicitly specified otherwise.

**Figure F.7:**
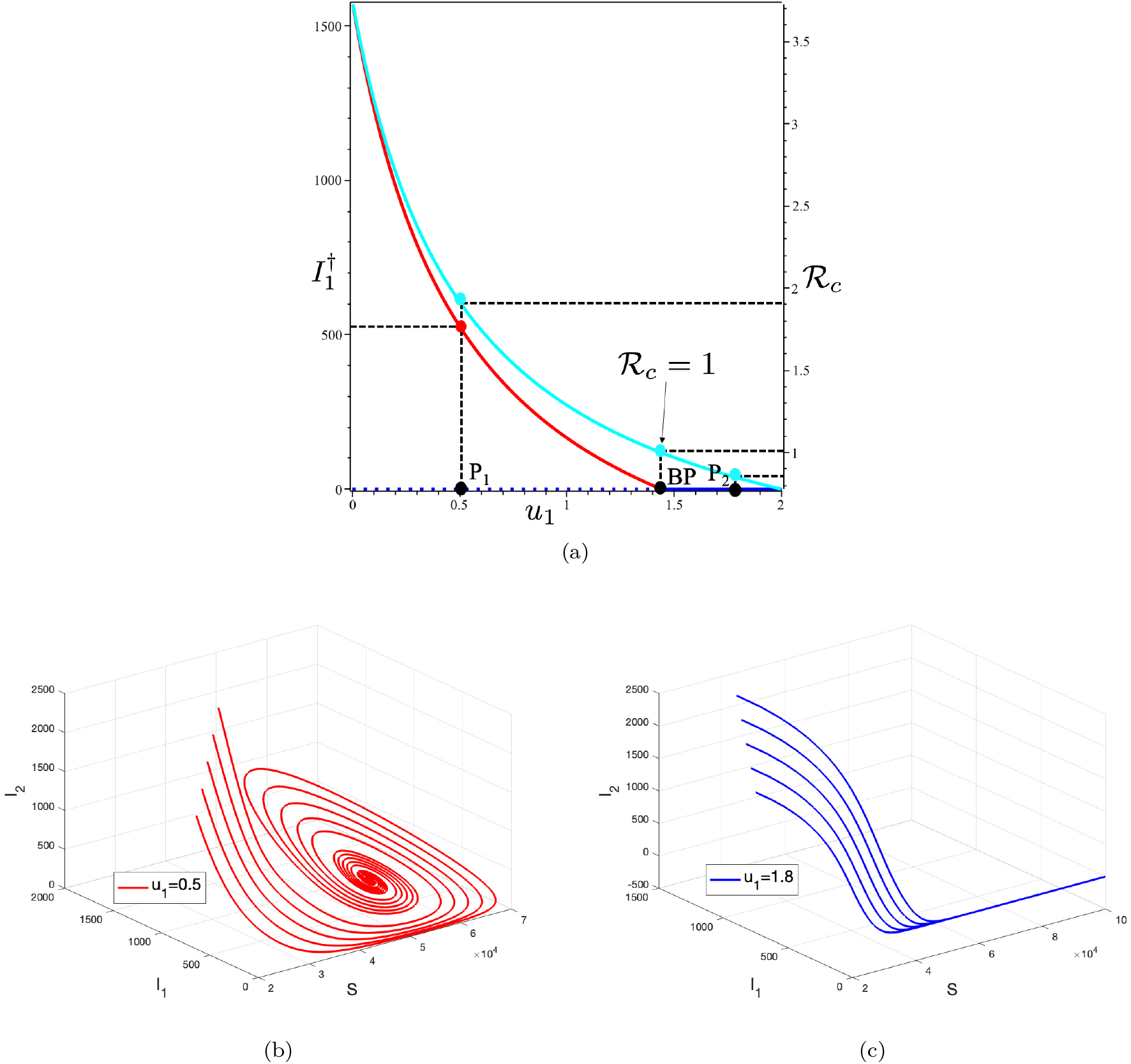
(a) Bifurcation diagram of system (1) represented by *I*_1_ and ℛ_*c*_ with respect to *u*_1_. The solid lines represent the stable branch of the equilibrium point and the dashed line represents the unstable branch of the equilibrium point. Red, blue and cyan curves represent the endemic equilibrium, TB-free equilibrium and ℛ_*c*_, respectively. Panel (b) and (c) show the trajectories in *S* − *I*_1_ − *I*_2_ phase planes at two different sample points *P*_1_(*u*_1_ = 0.5) and *P*_2_(*u*_1_ = 1.8), respectively. All solutions were obtained numerically using various initial conditions.

In Figure F.7, panel (a) illustrates the monotonic decrease of the endemic equilibrium (red curve) and the control reproduction number (cyan curve) with increasing of *u*_1_. The endemic equilibrium persists until reaching the Branching Point (BP) at ℛ_*c*_ = 1 (i.e., *u*_1_ = 1.438). Before the BP, the TB-free equilibrium is unstable (blue dotted line), while the TB-endemic equilibrium is stable (red solid curve). Once *u*_1_ surpasses the BP, the endemic equilibrium ceases to exist, and the stability of the TB-free equilibrium shifts from unstable to stable. To illustrate the impact of *u*_1_ on the dynamics of *I*_1_, *I*_2_ and *S*, we specifically selected two sample points, namely *P*_1_ (*u*_1_ = 0.5, ℛ_*c*_ = 1.916) and *P*_2_ (*u*_1_ = 1.8, ℛ_*c*_ = 0.844). Panel (b) depicts the solution of system (1) on the *S* − *I*_1_ − *I*_2_ plane for various initial conditions at sample point *P*_1_, while panel (c) provides the corresponding illustration for sample point *P*_2_.

**Figure F.8:**
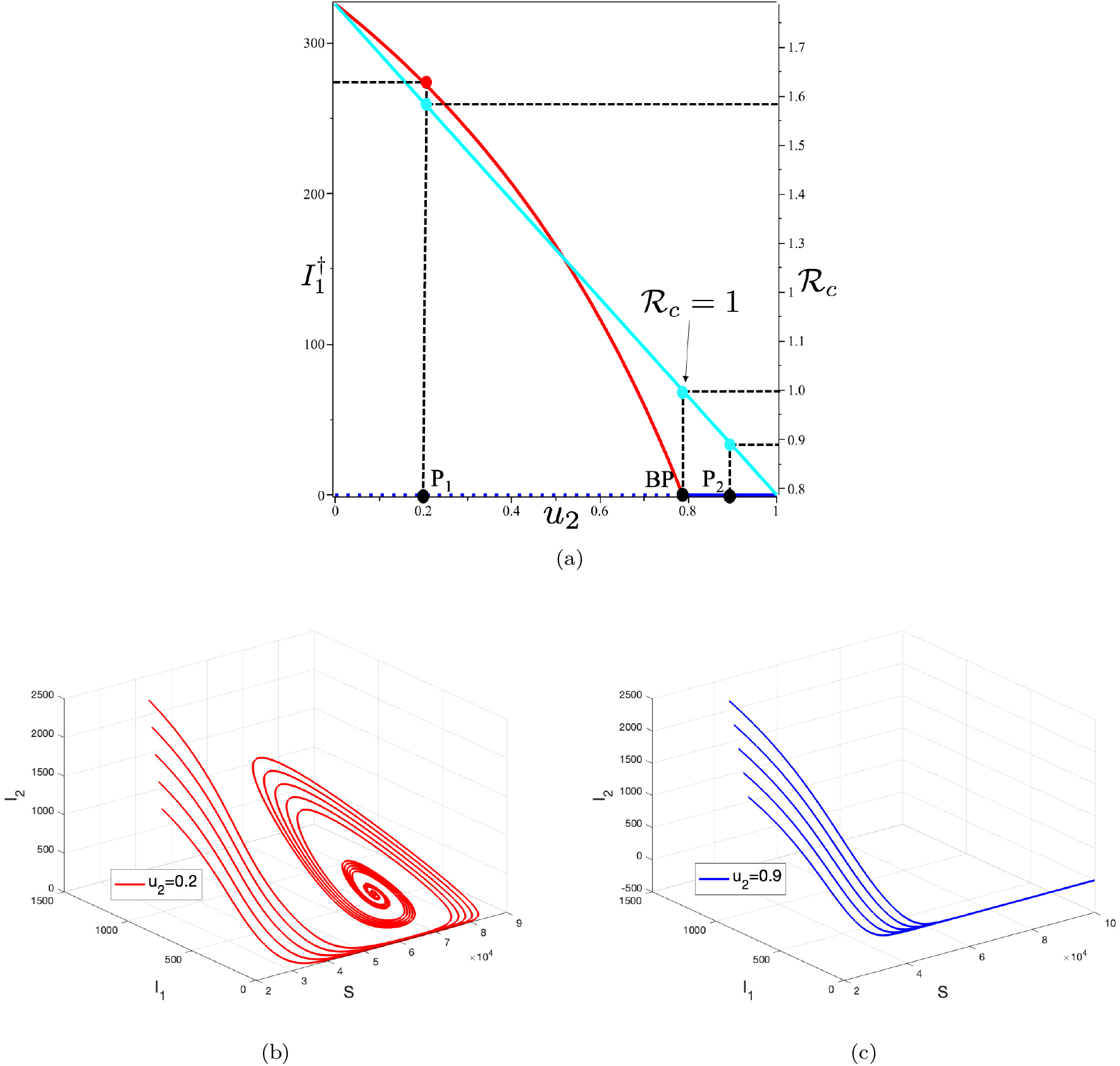
(a) Bifurcation diagram of system (1) represented by *I*_1_ and ℛ_*c*_ with respect to *u*_2_. The solid lines represent the stable branch of the equilibrium point and the dotted line represents the unstable branch of the equilibrium point. The red and cyan curves represent the endemic equilibrium and the ℛ_*c*_, respectively. Panel (b,c) shows of trajectories solution of *S, I*_1_ and *I*_2_ at two different sample points *P*_1_(*u*_2_ = 0.2) and *P*_2_(*u*_2_ = 0.9). All solutions were obtained numerically using various initial conditions.

The sensitivity of ℛ_*c*_ and equilibrium point size of *I*_1_ with respect to *u*_2_ is plotted in Figure F.8. We employed identical parameter values as those utilized for Figure F.7, with the exception of setting *u*_1_ = 1, while allowing *u*_2_ to remain a free parameter. In Panel (a), we can see that the intervention of *u*_2_ can significantly affect the ℛ_*c*_. Hence, a stable branch of TB-endemic equilibrium existed for *u*_2_ *<* 0.787, while an unstable branch was observed for *u*_2_ *>* 0.787. We selected two sample cases at points *P*_1_ and *P*_2_ with corresponding *u*_2_ values 0.2 and 0.9, respectively (see Figure F.8(a)), to illustrate the impacts of *u*_2_ on the dynamics of *I*_1_, *I*_2_ and *S*. At sample point *P*_1_, the solution converges to the endemic equilibrium point, while at *P*_2_, the solution converges towards the TB-free equilibrium point. Trajectories for both cases are displayed in Figures F.8, panels b and c, for various initial conditions.

For the last one-parameter sensitivity analysis, we show how ℛ_*c*_ and endemic size of *I*_1_ behave as *β* changes. It can be seen that larger *β* will increase ℛ_*c*_ and the endemic size of *I*_1_. The bifurcation diagram is shown in Figure F.9 panel (a). From the figure, we can see that the TB-free equilibrium is locally stable for *β <* 0.000011. As *β >* 0.000011, the stable branch of TB-free equilibrium point is bifurcated into an unstable TB-free equilibrium and a stable endemic equilibrium. The dynamics of *I*_1_ corresponding to sample points *P*_1_ and *P*_2_ are plotted in Figures F.9 panel (b) and (c), respectively. It is important to mention that the solutions tended to move towards the endemic equilibrium when *β* was larger than the BP, and the solutions converged towards the TB-free equilibrium when *β* was smaller than the BP.

**Figure F.9:**
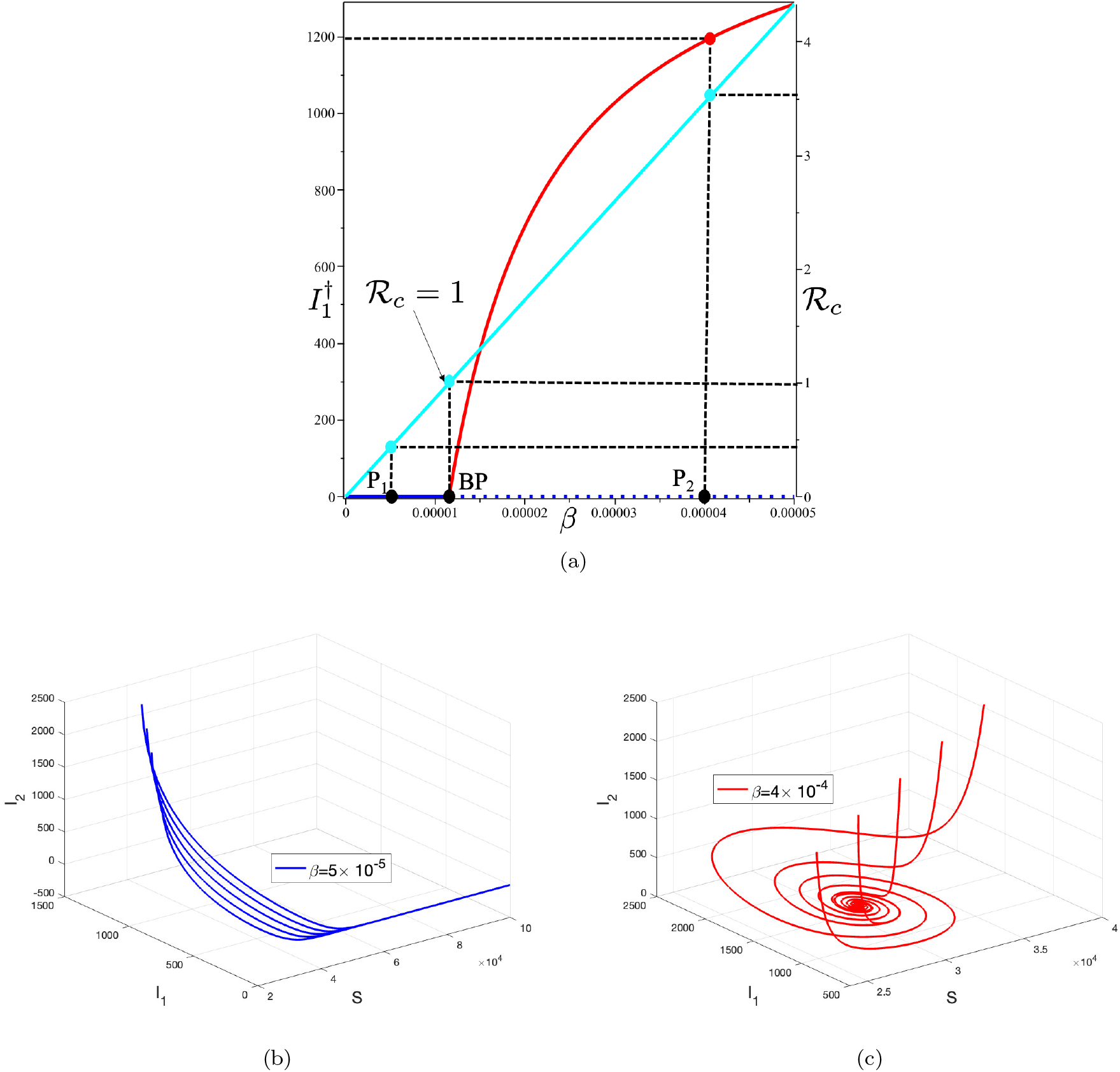
(a) Bifurcation diagram of system (1) represented by *I*_1_ and ℛ_*c*_ with respect to *β*. The red and cyan curves represent the endemic equilibrium and the ℛ_*c*_, respectively. Panel (b) and (c) show the trajectories of the solution for two different sample points *P*_1_(*β* = 0.000005) and *P*_6_(*β* = 0.00004). All solutions were obtained numerically using various initial conditions.

## Appendix G. Proof of Theorem (6): Characterization of the Optimal Control Problem

Let

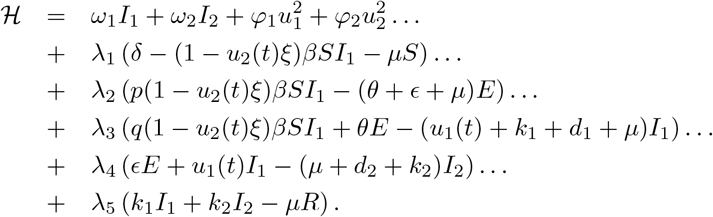

Differentiate Hamiltonian ℋ with respect to each control variables yields

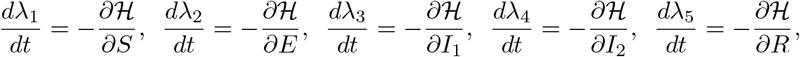

with it transversality condition *λ*_*i*_(*t*_*f*_) = 0 for *i* = 1, 2, 3, 4, 5.

The optimal condition of *u*_1_ taken by solving 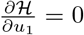 with respect to *u*_1_. Combine it with the lower and upper bound gives us

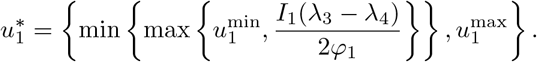

Similarly, we have

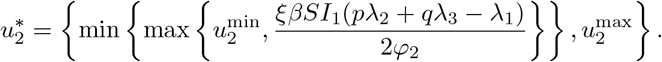

Hence the proof is completed.

## Appendix H. Forecasting of India, Lesotho, and Angola TB cases

**Figure H.10:**
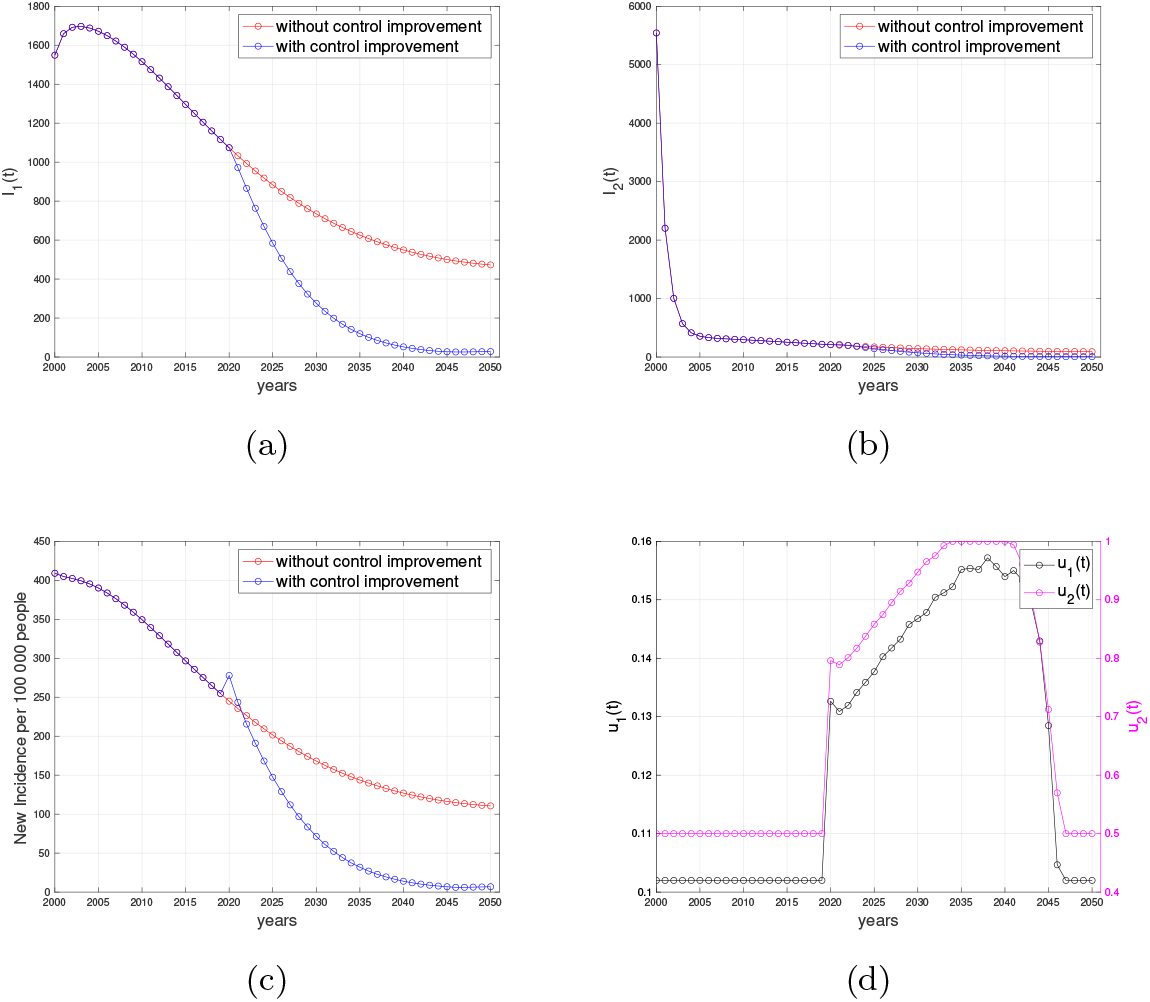
Forecasting and optimal control results for India data. Panel (a) and (b) represent the dynamic of *I*_1_ and *I*_2_, respectively. Panel (c) represent the case incidence per 100,000 people with and without control, while panel (d) shows the dynamic of controls.

**Figure H.11:**
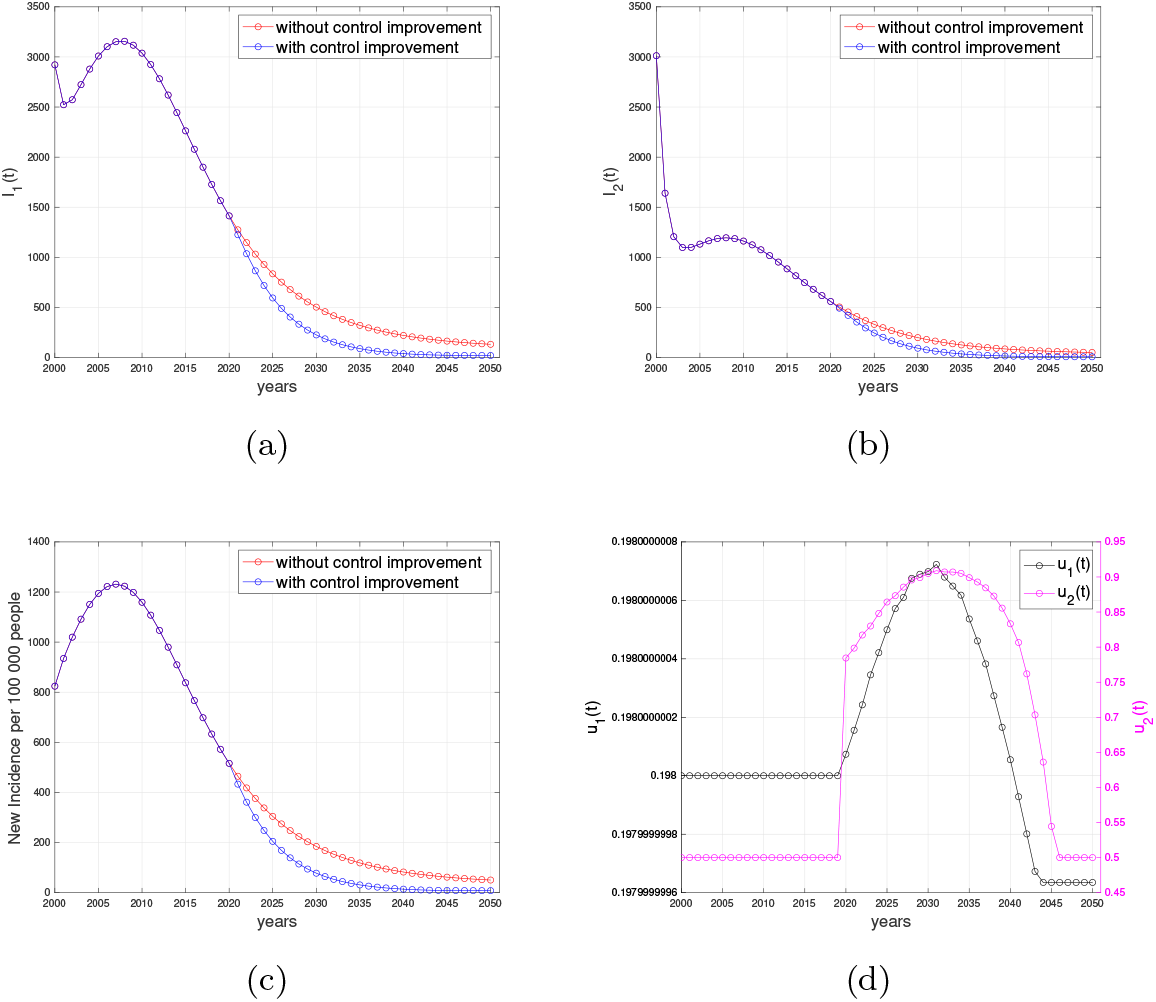
Forecasting and optimal control results for Lesotho data. Panel (a) and (b) represent the dynamic of *I*_1_ and *I*_2_, respectively. Panel (c) represent the case incidence per 100,000 people with and without control, while panel (d) shows the dynamic of controls.

**Figure H.12:**
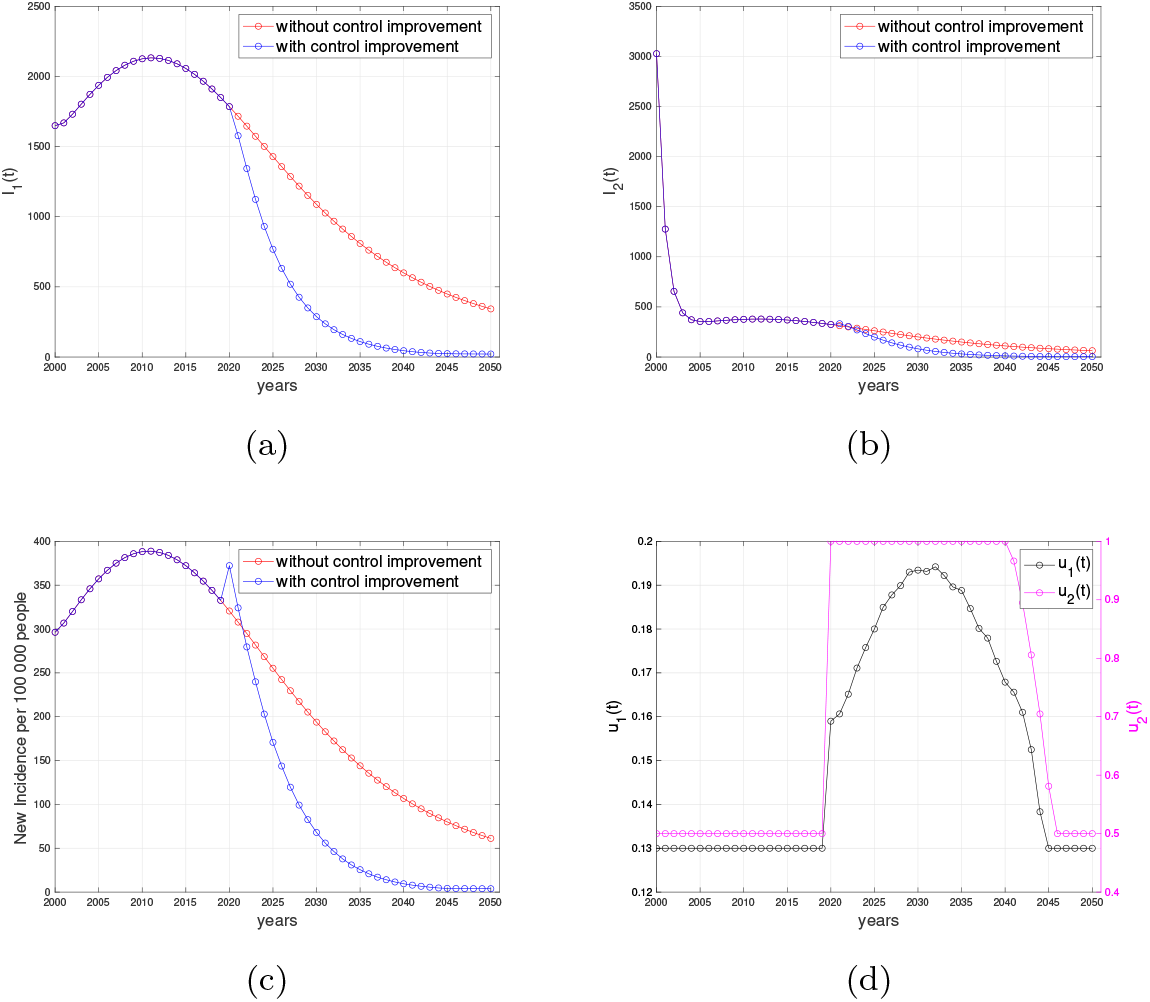
Forecasting and optimal control results for Angola data. Panel (a) and (b) represent the dynamic of *I*_1_ and *I*_2_, respectively. Panel (c) represent the case incidence per 100,000 people with and without control, while panel (d) shows the dynamic of controls.

## Appendix I. Numerical results on optimal control simulation for different strategy

**Figure I.13:**
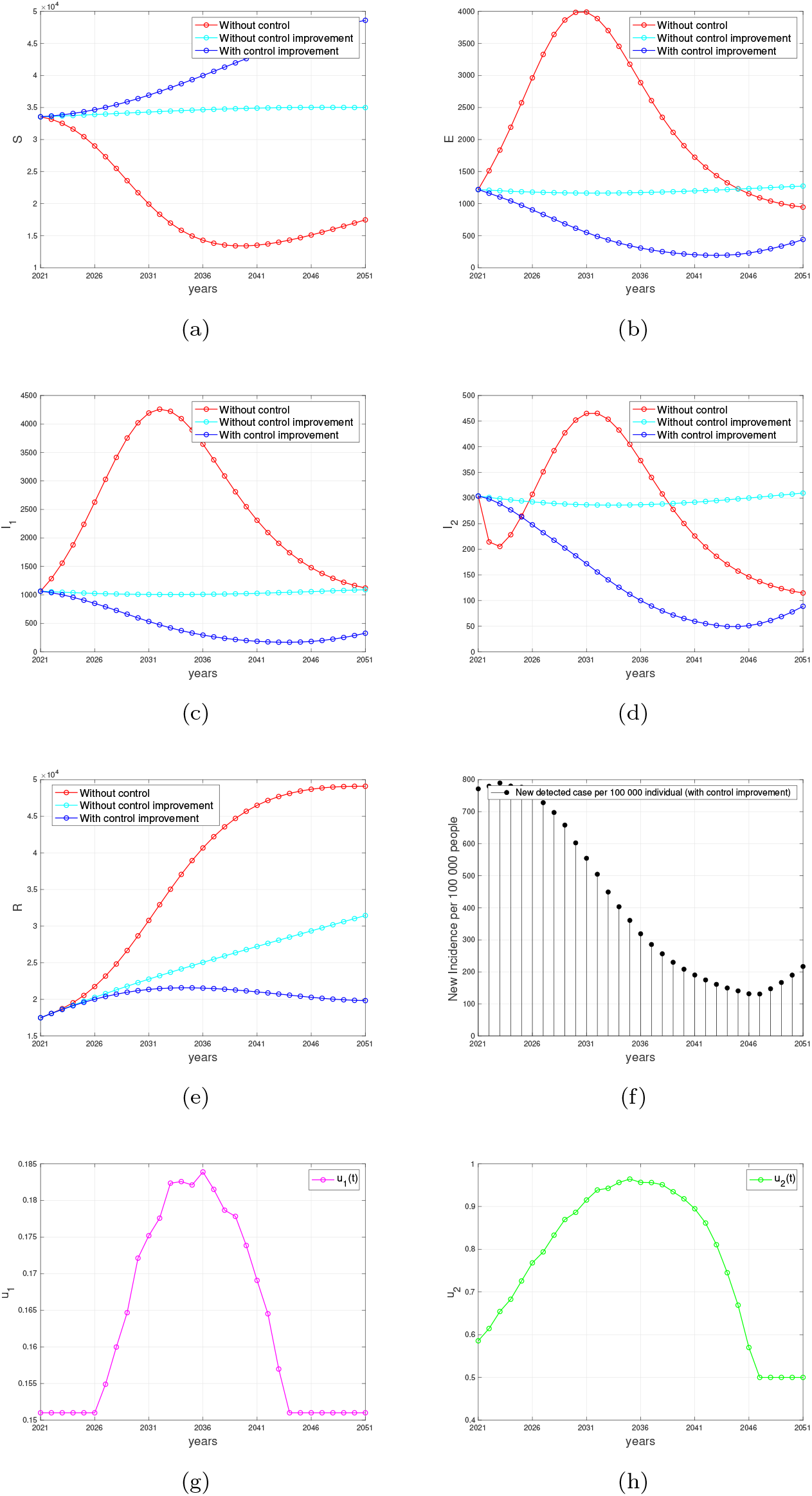
Forecasting and optimal control results for Indonesian data from 2021 to 2051 when case detection and medical mask use are implemented together. Panel (a) to (e) represent the dynamic of *S, E, I*_1_, *I*_2_, and *R*, respectively. Panel (f) represent the case incidence per 100,000 people while panel (g) and (h) shows the dynamic of control *u*_1_ and *u*_2_, respectively.

**Figure I.14:**
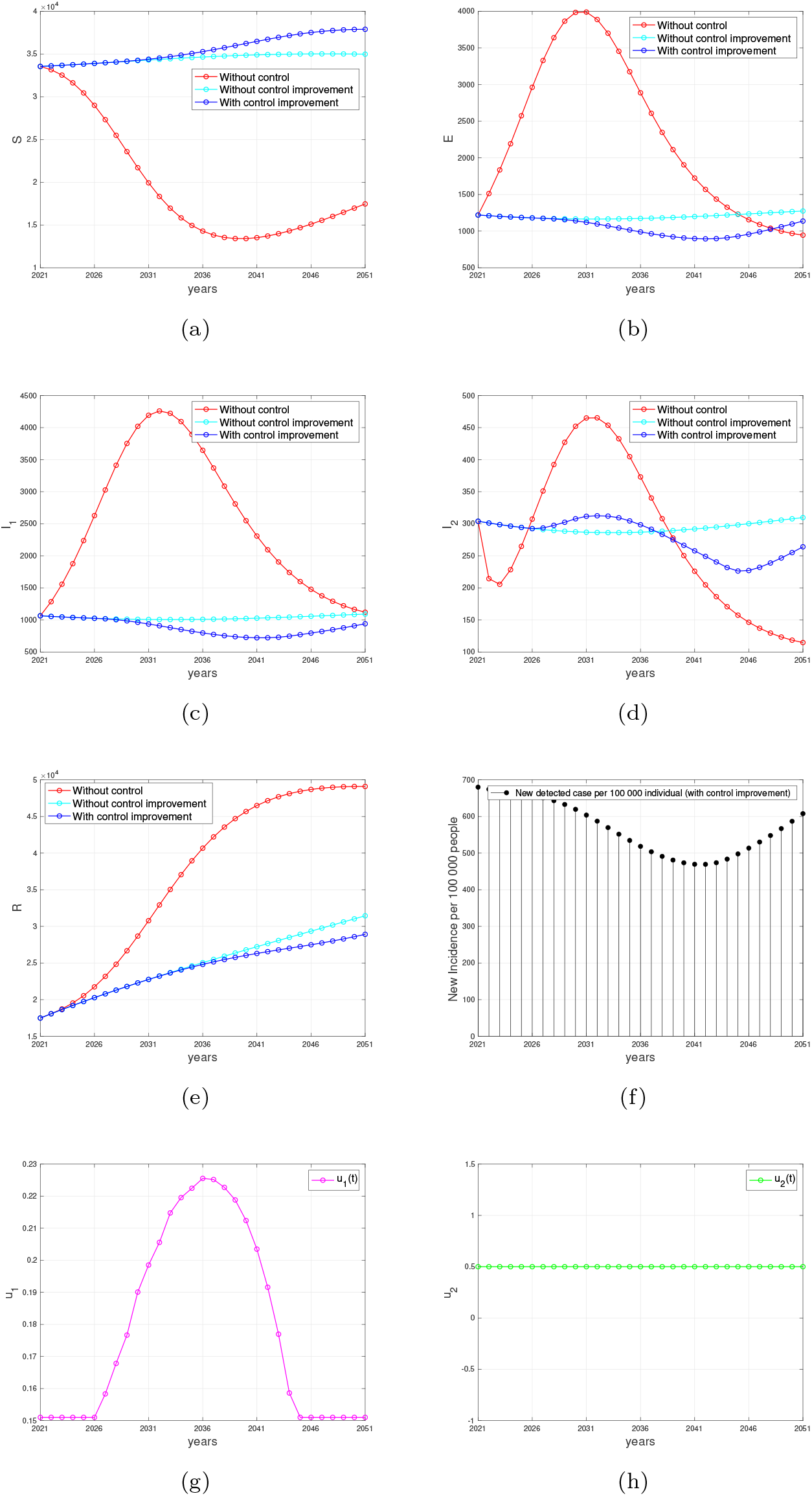
Forecasting and optimal control results for Indonesian data from 2021 to 2051 when case detection intervention improved, but medical mask left as a constant at *u*_2_ = 0.5. Panel (a) to (e) represent the dynamic of *S, E, I*_1_, *I*_2_, and *R*, respectively. Panel (f) represent the case incidence per 100,000 people while panel (g) and (h) show the dynamic of control *u*_1_ and *u*_2_, respectively.

**Figure I.15:**
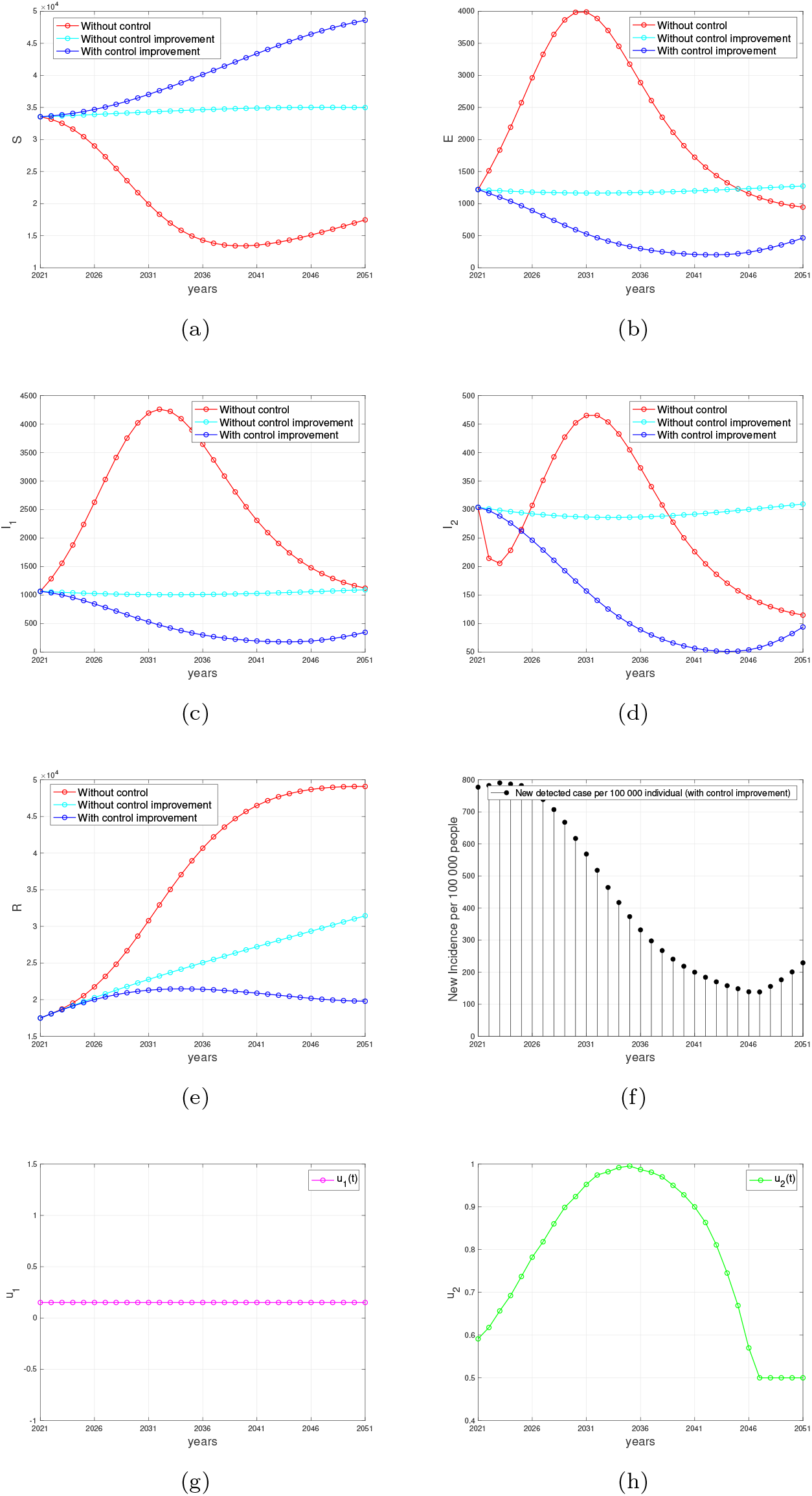
Forecasting and optimal control results for Indonesian data from 2021 to 2070 when medical mask intervention improved, but case detection left as a constant at *u*_1_ = 0.151. Panel (a) to (e) represent the dynamic of *S, E, I*_1_, *I*_2_, and *R*, respectively. Panel (f) represent the case incidence per 100 000 people while panel (g) and (h) shows the dynamic of control *u*_1_ and *u*_2_, respectively.

